# Association of Inflammation with Depression and Anxiety: Evidence for Symptom-Specificity and Potential Causality from UK Biobank and NESDA Cohorts

**DOI:** 10.1101/2021.01.08.20248710

**Authors:** Yuri Milaneschi, Nils Kappelmann, Zheng Ye, Femke Lamers, Sylvain Moser, Peter B. Jones, Stephen Burgess, Brenda W. J. H. Penninx, Golam M. Khandaker

## Abstract

We examined whether inflammation is uniformly associated with all depressive and anxiety symptoms, and whether these associations are potentially causal. Data was from 147,478 individuals from the UK Biobank (UKB) and 2,905 from the Netherlands Study of Depression and Anxiety (NESDA). Circulating C-reactive protein (CRP) was measured in both cohorts and interleukin-6 (IL-6) in NESDA. Genetic instruments for these proteins were obtained from published GWAS and UKB. Depressive and anxiety symptoms were assessed with self-report questionnaires. In NESDA, neurovegetative (appetite, sleep, psychomotor) symptoms were disaggregated as increased vs. decreased. In joint analyses, circulating CRP was associated with depressive symptoms of depressed mood (OR=1.06, 95%CI=1.05-1.08), altered appetite (OR=1.25, 95%CI=1.23-1.28), sleep problems (OR=1.05, 95%CI=1.04-1.06), and fatigue (OR=1.12, 95%CI=1.11-1.14), and with anxiety symptoms of irritability (OR=1.06, 95%CI=1.05-1.08) and worrying control (OR=1.03, 95%CI=1.02-1.04). Further analyses in NESDA using IL-6 as exposure confirmed associations with depressive symptoms, including anhedonia (OR=1.30, 95%CI=1.12-1.52). Both CRP (OR=1.27, 95%CI=1.13-1.43) and IL-6 (OR=1.26, 95%CI=1.07-1.49) were associated with increased sleep. CRP was associated with increased appetite (OR=1.21, 95%CI=1.08-1.35) while IL-6 with decreased appetite (OR=1.45, 95%CI=1.18-1.79). In Mendelian Randomization analyses, increased risk of fatigue (estimate=0.25, SE=0.08) and sleep problems (estimate=0.19, SE=0.07) were associated with genetically-predicted higher IL-6 activity. Inflammation was associated with core depressive symptoms of low mood and anhedonia and somatic/neurovegetative symptoms of fatigue, altered sleep and appetite changes. Less consistent associations were found for anxiety. The IL-6/IL-6R pathway could be causally linked to depression. Experimental studies are required to further evaluate causality, mechanisms, and usefulness of immunotherapies for depressive symptoms.

## INTRODUCTION

A role for inflammatory dysregulation in depression has been suggested by a large body of evidence. Clinical studies have shown that a quarter of patients with hepatitis C develop a depressive episode following pro-inflammatory interferon treatment [1]. Large meta-analyses report cross-sectional and longitudinal associations between inflammatory markers - such as C-Reactive Protein (CRP) and Interleukin 6 (IL-6) - and depression [2–7]. Indeed, inflammation is present in about a quarter of depressed patients as evidenced by elevated CRP levels [8]. However, there are key outstanding questions regarding symptom specificity and potential causality of association.

Previous studies have mainly used composite measures of depression, but it is a phenotypically heterogenous syndrome. Depression is also highly comorbid with anxiety, but studies of inflammation and anxiety symptoms are scarce [9]. It is possible that inflammation is relevant for some but not all affective symptoms. Higher inflammatory marker levels have been reported to be mainly associated with anhedonia and neurovegetative symptoms such as fatigue, appetite and sleep alterations [10–16]. However, assessments in previous research often conflated divergent alterations in neurovegetative symptoms (e.g. increased vs decreased appetite). Emerging evidence suggests that inflammatory and metabolic alterations map more consistently onto “atypical” energy-related symptoms, particularly increased sleep, appetite/weight, fatigue and leaden paralysis [17–19]. Inflammation is unlikely to be relevant for all cases of depression. A symptom-based approach may provide insights into mechanisms of inflammation-related depression and could help inform patient selection in immunotherapy trials.

In addition to symptom-specificity, another key issue is causality of association. Mendelian Randomization (MR) is an approach to evaluate potential causality, which uses genetic variants as proxy instruments (unrelated to confounding variables based on Mendel’s law of random allele segregation) to test exposure-outcome associations.[20] A recent MR study of depressive symptoms indicated that IL-6 signalling could be causally linked with suicidality [21]. However, this study did not investigate anxiety symptoms or particular direction of change (increase vs decrease) in neurovegetative symptoms specifically. MR studies of depressive and anxiety symptoms including more granular information on direction of symptom change is required to gain greater insights into the potential role of inflammation in these disorders.

We have examined specificity and potential causality of associations for CRP and IL-6 with depressive and anxiety symptoms using large-scale data from two well-established European cohorts, UK Biobank (UKB) and Netherlands Study of Depression and Anxiety (NESDA). In addition to testing symptom-level associations for CRP in two cohorts, we have carried out further analysis in the NESDA cohort using: (1) IL-6 levels as exposure; and (2) granular information on direction of change for particular symptoms (e.g., sleep problems, appetite alterations) as outcomes in relation to IL-6 and CRP. Furthermore, we have carried out one- and two-sample MR analyses to test whether associations of IL-6 and CRP with specific depressive and anxiety symptoms are likely to be causal using genetic variants regulating levels/activity of these inflammatory markers as instrumental variables.

## METHODS

### Study cohorts

The present study utilised data from the UK Biobank (UKB) study [22], a population-based cohort comprising 502,524 UK residents aged 40-69 years, and the Netherlands Study of Depression and Anxiety (NESDA) [23], an ongoing cohort study of 2,981 participants aged 18-65 years with current or past depressive and/or anxiety disorder and healthy controls. Detailed descriptions of study cohorts are available as Supplementary Methods.

Briefly, we included up to 147,478 participants from the UKB sample with data on CRP and depressive/anxiety symptoms. For Mendelian Randomisation analysis, a sample of unrelated individuals with European ancestry and genotype information was used including up to 325,441 participants to estimate single-nucleotide polymorphism (SNP)-exposure associations with CRP for one-sample MR analysis and up to 111,572 participants to estimate SNP-outcome associations. From the NESDA sample, 2,905 participants with complete data at baseline on inflammatory markers and depressive/anxiety symptoms were selected. Biomarkers and symptoms were assessed again at 2-year and 6-year follow-up, totalling ∼7,000 observations.

The UKB study was approved by the North West Centre for Research Ethics Committee and Human Tissue Authority research tissue bank. The current analysis was approved under project no. 26999. The NESDA research protocol was approved centrally by the Vrije Universiteit (VU) Amsterdam University Medical Centre ethics committee and locally by the ethics committee of participating universities. Participants from both cohorts provided informed consent.

### Depressive and anxiety symptoms

In UKB nine depressive and seven anxiety symptoms were assessed using Patient Health Questionnaire (PHQ)-9 and Generalized Anxiety Disorder (GAD)-7 questionnaires, respectively [24]. We identified similar items from NESDA that were assessed by different questionnaires (i.e., Inventory of Depressive Symptomatology [IDS-SR_30_], Beck Anxiety Inventory [BAI] & Penn State Worry Questionnaire [PSWQ]) at baseline, 2- and 6-year follow-up. Items coding similar domains (e.g., anhedonia assessed as lack of “general interest” and “capacity for pleasure”) and items coding specific neurovegetative symptoms (e.g., sleeping problems separately measured as “increased” or “decreased” sleep) were conflated to align these with UKB data for ease of comparison. All symptoms were binarised to reflect a measure of any versus no symptom endorsement. For depressive symptoms we also created two summary scores, “psychological” and “somatic”, based on a 2-factor model based on reported genetic covariance among nine PHQ-9 symptoms in UKB [25].

In extended NESDA analyses, items measuring neurovegetative symptoms were left disaggregated coding for specific alterations. Details on questionnaires, items and coding are in Supplementary Methods and Supplementary Table 1.

### Inflammatory markers

In both UKB and NESDA, circulating CRP levels were measured using high-sensitivity assays. In NESDA, circulating IL-6 levels were additionally available, and both inflammatory markers were assessed at baseline, 2- and 6-year follow-up which were modelled in repeated-measurement analyses. Details on blood sampling and technical assay features are described in Supplementary Methods.

### Covariates

The same set of covariates was considered in both cohorts, which included age, sex, socioeconomic status (SES), smoking, alcohol consumption, physical activity, history of type two diabetes (T2D) and cardiovascular diseases (CVD), and Body Mass Index (BMI). SES was measured via the Townsend Deprivation Index (a composite score of deprivation derived from national census data [26]) in UKB, and as years of education in NESDA. Covariate measurements and their distribution are described in Supplementary Methods and Supplementary Tables 2 and 3.

### Statistical Analysis

#### Cross-cohort analyses of associations between CRP and depressive/anxiety symptoms

Associations between CRP levels and depressive/anxiety symptoms were estimated by regressing individual symptoms and summary scores on log-transformed values of CRP. Details of statistical models are given in Supplementary Methods. In order to explore the impact of covariates on the association between CRP and symptoms, estimates were adjusted for age, sex and SES (Model 1), additionally adjusted for smoking, alcohol consumption, physical activity and T2D/CVD (Model 2), and additionally adjusted for BMI (Model 3).

Cohort-specific estimates were pooled using random-effects meta-analysis with the DerSimonian and Laird method [27]. For each model, False-Discovery Rate (FDR) q-values were calculated taking into account testing across 16 symptoms and 2 summary scales.

#### Extended analyses using IL-6 levels and additional symptoms in NESDA

Items measuring neurovegetative (appetite, sleep and psychomotor) symptoms were disaggregated in order to estimate associations between specific alterations (e.g., increase vs decrease) and inflammation. We also used (log)IL-6 levels as exposure, not measured in UKB. For each model, FDR q-values were calculated accounting for testing across 21 symptoms and two summary scales.

### Mendelian Randomization analyses

#### Instrument selection

We used multiple genetic instruments for CRP [28, 29], based on SNPs in the *CRP* gene associated with serum CRP concentrations, and for IL-6 [28, 30, 31], based on SNPs in the *IL-6R* gene associated either with serum CRP concentrations (as downstream readout of IL-6) or with serum IL-6 concentrations. Further details are provided as Supplementary Methods and in Supplementary Table 4.

#### MR analysis

Availability of CRP concentrations in the UKB sample allowed both one-sample and two-sample MR analyses with SNP-exposure estimates obtained from original reports (two-sample MR) or by regressing CRP on SNPs in UKB (one-sample MR). SNP-outcome estimates were all obtained by regressing outcome phenotypes on SNPs. Regression analyses were controlled for 20 genotype principal components, age, age^2^, sex, and age*sex. We performed standard variant harmonisation procedures on obtained estimates [32]. As main analysis, we used fixed-effects inverse variance weighted (IVW) meta-analysis per exposure-outcome combination or Wald ratio estimation for the single-SNP IL-6 instrument [30]. Potential horizontal pleiotropy was evaluated using Cochrane’s *Q* statistic [33]. Further details are noted in Supplementary Methods.

## RESULTS

Table 1 shows main variables including symptom endorsement and inflammatory marker levels in UKB and NESDA cohorts.

**Table 1.** Main variables of interest in the two cohorts.

|  | **UK Biobank**<br>*N = 143,465* | **NESDA**<br>*N = 2,905* |
| --- | --- | --- |
| *Sociodemographics* |  |  |
| **Age years (mean ± SD)** | 55.9 (7.7) | 41.9 (13.1) |
| **Sex (F) (%)** | 56.2 | 66.5 |
| *Symptom endorsement - Depression (%)* |  |  |
| **D.1 Anhedonia** | 18.5 | 13.9 |
| **D.2 Depressed mood** | 21.9 | 21.6 |
| **D.3 Sleep problem** | 48.7 | 25.9 |
| **D.4 Fatigues** | 49.9 | 36.5 |
| **D.5 Appetite change** | 18.2 | 7.7 |
| **D.6 Feelings of inadequacy** | 19.2 | 24.2 |
| **D.7 Cognitive problems** | 17.9 | 21.9 |
| **D.8 Psychomotor change** | 5.5 | 18.1 |
| **D.9 Suicidal ideation** | 4.3 | 12.2 |
| *Symptom summary scales* |  |  |
| **Psychological symptoms (mean ± SD)** | 0.74 (1.51) | 0.721 (1.04) |
| **Somatic symptoms (mean ± SD)** | 1.65 (2.03) | 1.09 (1.30) |
| *Symptom endorsement - Anxiety (%)* |  |  |
| **A.1 Anxiety** | 28.0 | 24.5 |
| **A.2 Worrying control** | 23.4 | 24.8 |
| **A.3 Generalized worrying** | 31.6 | 31.5 |
| **A.4 Lack of relaxation** | 28.3 | 32.3 |
| **A.5 Restlessness** | 11.8 | 25.7 |
| **A.6 Irritability** | 27.2 | 21.0 |
| **A.7 Foreboding** | 16.6 | 20.4 |
| *Inflammatory markers* |  |  |
| **CRP (mg/L) (median, IQR)** | 1.15 (0.58-2.37) | 1.22 (0.54 - 3.00) |
| **IL-6 (pg/mL) (median, IQR)** | NA | 0.75 (0.49 - 1.25) |

*Note:* UK Biobank sample size reflects all individuals with complete symptom and CRP data. For NESDA baseline values are reported; measures at 2- and 6-year follow-up are reported in Supplementary Table 5 and Supplementary Figures 1 & 2. NA=not applicable.

### Association between CRP and depressive/anxiety symptoms in UKB and NESDA

Figure 1 shows results from individual cohorts and Table 2 shows meta-analytic pooled estimates representing the associations of CRP with individual depressive/anxiety symptoms and summary scores for somatic and psychological symptoms of depression (exact numbers in Supplementary Tables 6 & 7). After adjustment for sociodemographic, lifestyle and health-related factors (Model 2), higher CRP was associated with appetite change (OR=1.25, 95%CI=1.23-1.28), fatigue (OR=1.12, 95%CI=1.11-1.14), depressed mood (OR=1.06, 95%CI=1.05-1.08), and sleep problems (OR=1.05, 95%CI=1.04-1.06) among depressive symptoms, and with irritability (OR=1.06, 95%CI=1.05-1.08) and worrying control (OR=1.03, 95%CI=1.02-1.04) among anxiety symptoms. CRP was not associated with somatic and psychological symptom summary scores for depression. Evidence for associations attenuated with increasing confounder adjustment and especially after including BMI; however, associations of CRP with fatigue, sleep problems, depressed mood and irritability remained statistically significant.

**Table 2.** Pooled Association Results between CRP and Depressive/Anxiety Symptoms.

| Symptom | Model 1 |  |  | Model 2 |  |  | Model 3 |  |  |
| --- | --- | --- | --- | --- | --- | --- | --- | --- | --- |
|  | OR (95% CI) | P | Q | OR (95% CI) | P | Q | OR (95% CI) | P | Q |
| *Depressive symptoms* |  |  |  |  |  |  |  |  |  |
| Anhedonia | 1.13 (1.03-1.23) | 0.01 | 0.02 | 1.08 (0.99-1.18) | 0.101 | 0.203 | 1.03 (0.96-1.11) | 0.423 | 0.789 |
| Depressed mood | 1.1 (1.07-1.13) | <0.001 | <0.001 | 1.06 (1.05-1.08) | <0.001 | <0.001 | 1.03 (1.01-1.04) | <0.001 | 0.002 |
| Sleeping problems | 1.07 (1.06-1.08) | <0.001 | <0.001 | 1.05 (1.04-1.06) | <0.001 | <0.001 | 1.02 (1.01-1.03) | <0.001 | 0.003 |
| Fatigue | 1.17 (1.15-1.18) | <0.001 | <0.001 | 1.12 (1.11-1.14) | <0.001 | <0.001 | 1.06 (1.05-1.07) | <0.001 | <0.001 |
| Appetite changes | 1.3 (1.2-1.4) | <0.001 | <0.001 | 1.25 (1.23-1.28) | <0.001 | <0.001 | 1.02 (0.96-1.1) | 0.499 | 0.789 |
| Feelings of inadequacy | 1.02 (0.91-1.14) | 0.734 | 0.777 | 1 (0.9-1.1) | 0.925 | 0.979 | 0.98 (0.92-1.05) | 0.564 | 0.789 |
| Cognitive problems | 1.08 (1.01-1.16) | 0.034 | 0.055 | 1.05 (0.99-1.11) | 0.092 | 0.203 | 1.02 (1-1.03) | 0.043 | 0.128 |
| Psychomotor changes | 1.07 (0.87-1.3) | 0.537 | 0.644 | 1.03 (0.86-1.22) | 0.778 | 0.875 | 0.99 (0.86-1.13) | 0.859 | 0.859 |
| Suicidal ideation | 1.04 (0.86-1.26) | 0.683 | 0.768 | 1 (0.86-1.17) | 0.991 | 0.991 | 0.99 (0.91-1.09) | 0.856 | 0.859 |
| *Anxiety symptoms* |  |  |  |  |  |  |  |  |  |
| Anxiety | 1.01 (1-1.03) | 0.012 | 0.022 | 1 (0.99-1.01) | 0.758 | 0.875 | 1 (0.99-1.02) | 0.606 | 0.789 |
| Worrying control | 1.05 (1.03-1.08) | <0.001 | <0.001 | 1.03 (1.02-1.04) | <0.001 | <0.001 | 1.01 (1-1.03) | 0.033 | 0.12 |
| Generalised worrying | 1.03 (1.01-1.04) | <0.001 | <0.001 | 1.01 (0.99-1.02) | 0.309 | 0.505 | 1 (0.99-1.01) | 0.833 | 0.859 |
| Lack of relaxation | 1.04 (1.02-1.05) | <0.001 | <0.001 | 1.01 (1-1.02) | 0.057 | 0.147 | 1.01 (1-1.02) | 0.206 | 0.464 |
| Restlessness | 0.98 (0.84-1.15) | 0.834 | 0.834 | 0.96 (0.85-1.1) | 0.586 | 0.812 | 0.96 (0.86-1.08) | 0.52 | 0.789 |
| Irritability | 1.09 (1.07-1.1) | <0.001 | <0.001 | 1.06 (1.05-1.08) | <0.001 | <0.001 | 1.03 (1.01-1.04) | <0.001 | <0.001 |
| Foreboding | 1.04 (0.98-1.1) | 0.21 | 0.29 | 1.01 (0.96-1.07) | 0.677 | 0.871 | 1 (0.97-1.02) | 0.812 | 0.859 |
| *Depressive symptom score* | **β (SE)** | **P** | **Q** | **β (SE)** | **P** | **Q** | **β (SE)** | **P** | **Q** |
| Psychological symptoms | 0.043 (0.042) | 0.312 | 0.402 | 0.023 (0.029) | 0.42 | 0.63 | 0.006 (0.012) | 0.614 | 0.789 |
| Somatic symptoms | 0.14 (0.11) | 0.203 | 0.29 | 0.099 (0.077) | 0.2 | 0.36 | 0.041 (0.028) | 0.144 | 0.371 |

**Figure 1.**
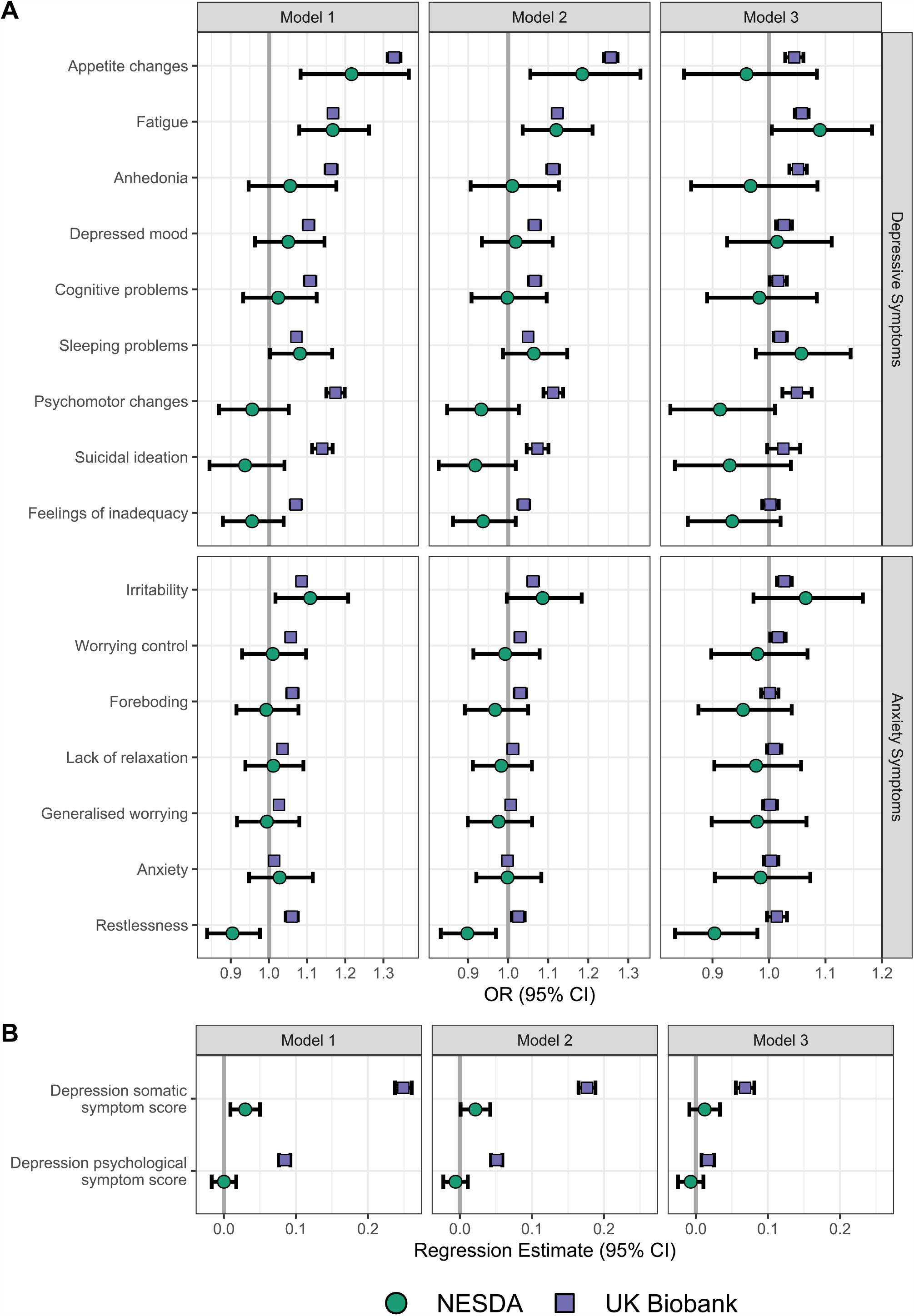
Association estimates of CRP with depressive and anxiety symptoms from UKB and NESDA cohorts. Association estimates are shown with individual depressive and anxiety symptoms (A) and depressive summary scores (B). Models have been adjusted for age, sex and SES (Model 1), additionally adjusted for smoking, alcohol consumption, physical activity and T2D/CVD (Model 2), and additionally adjusted for BMI (Model 3).

### Further analyses using IL-6 levels and additional symptoms in NESDA

Among symptoms identified in cross-cohort analysis, extended analyses examining (log)IL-6 levels and disaggregated neurovegetative symptoms confirmed associations with altered sleep, appetite and fatigue (Figure 2, full results Supplementary Tables 8 & 9). In particular, both higher CRP and IL-6 showed converging associations with hypersomnia (CRP OR=1.27, 95%CI=1.13-1.43; IL-6 OR=1.26, 95%CI=1.07-1.49) and fatigue (CRP OR=1.12, 95%CI=1.04-1.21; IL-6 OR=1.19, 95%CI=1.07-1.33). In contrast, divergent associations with appetite alterations emerged, with CRP linked to increased appetite (OR=1.21, 95%CI=1.08-1.35) and IL-6 linked to decreased appetite (OR=1.45, 95%CI=1.18-1.79). IL-6 but not CRP was also associated with anhedonia (OR=1.30, 95%CI=1.12-1.52). Additional adjustment for BMI did not change results substantially, but the association between CRP and increased appetite was no longer significant. In model 2, higher IL-6 was associated with depressed mood, but not after considering multiple testing. IL-6 was not associated with anxiety symptoms.

**Figure 2.**
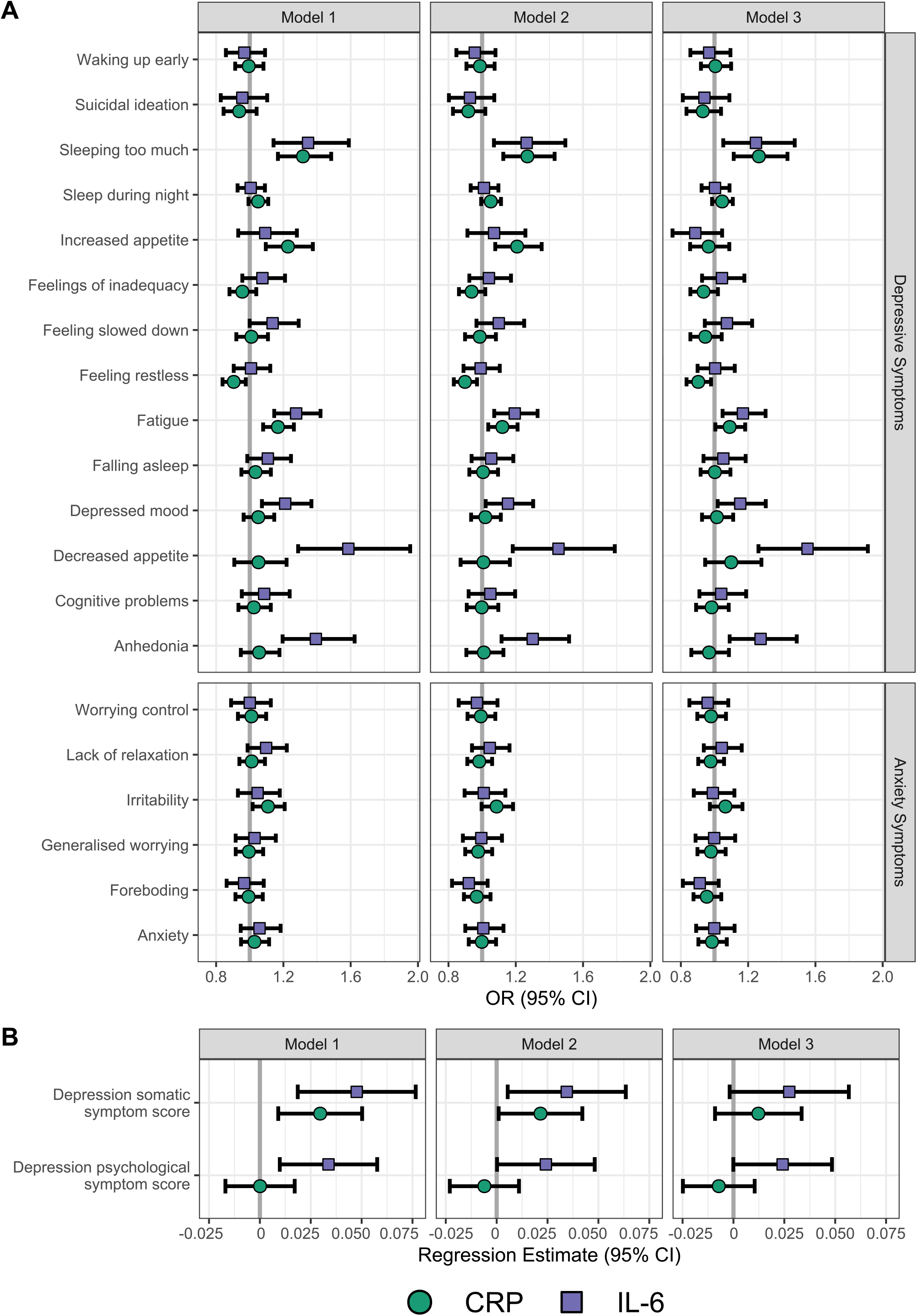
NESDA association estimates of CRP and IL-6 with depressive and anxiety symptoms. Association estimates are shown with individual depressive and anxiety symptoms (A) and depressive summary scores (B). Models have been adjusted for age, sex and SES (Model 1), additionally adjusted for smoking, alcohol consumption, physical activity and T2D/CVD (Model 2), and additionally adjusted for BMI (Model 3).

### Results for Mendelian randomization analyses

IVW one-sample and two-sample MR results for CRP and IL-6 instruments are displayed in Figure 3 based on genetic instruments derived from Georgakis *et al*.[28] Exact numeric results for these and other instruments are provided in Supplementary Tables 10 and 11.

**Figure 3.**
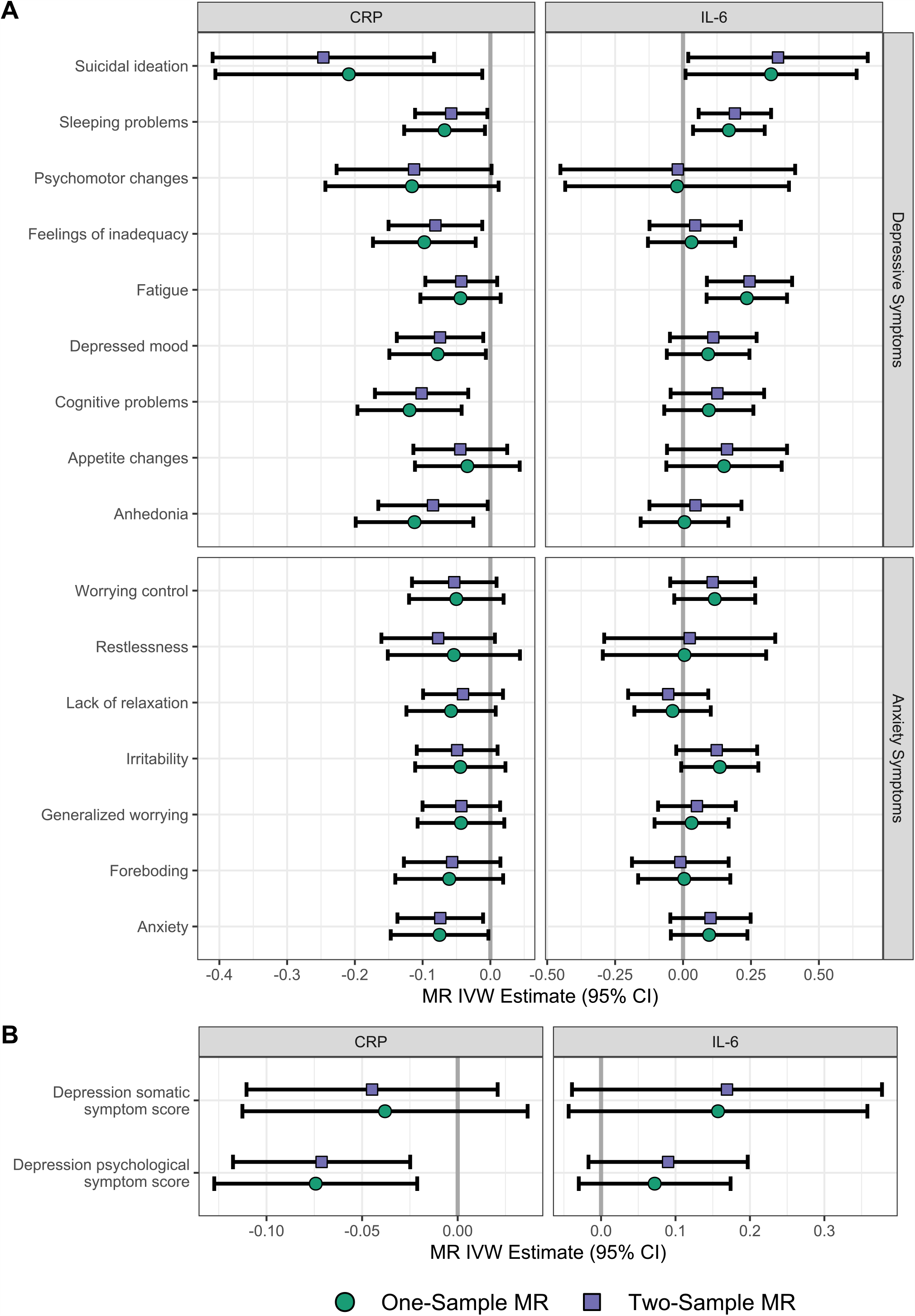
Mendelian randomisation results of CRP and IL-6 with depressive and anxiety symptoms. MR association estimates are shown with individual depressive and anxiety estimates (A) and depression summary scores (B). Results reflect MR IVW estimates based on Georgakis *et al*.[28] instruments; exact numeric values are presented in Supplementary Tables 11 & 12.

Overall, MR results showed that genetically predicted higher CRP levels were associated with lower risk of depressive and anxiety symptoms. After adjusting for multiple comparisons, evidence for associations remained for suicidal ideation, cognitive problems, and the psychological symptom summary score.

On the other hand, MR results for IL-6 showed that genetically predicted increased IL-6 signalling was associated with increased fatigue and sleep problems, which persisted after corrections for multiple testing. We also replicated a previously reported MR association between IL-6 and suicidal ideation (cf. Kappelmann *et al*.[21]), but this association did not survive multiple comparison correction.

Assessment of heterogeneity did not indicate that the aforementioned associations were likely to be due to horizontal pleiotropy except for the MR associations between lower CRP with increased suicidality (Supplementary Table 12 & 13).

## DISCUSSION

Using large-scale data from two well-established cohorts, we report an extensive evaluation of the associations between inflammatory markers and individual symptoms of depression and anxiety, including cross-cohort analyses, extended phenotype analyses on symptoms assessed at more granular resolutions, and MR analysis testing potential causality. Our results provided evidence for symptom-specificity. Inflammation does not map uniformly onto all affective symptoms, but rather are more consistently associated with specific depressive symptoms of fatigue, altered appetite, sleep problems (in particular hypersomnia), and the core symptom of depressed mood as compared to other symptoms of depression and anxiety. Furthermore, we provide evidence consistent with a potentially causal role of IL-6 in fatigue and sleep alterations; please see Supplementary Table 15 for an overview of main results.

Results across different analytical models and biomarkers highlighted more consistent associations of inflammation with depressive than with anxiety symptoms. For anxiety, associations were mainly limited to CRP and irritability, a symptom also commonly present in depression. These results align with an extensive evidence-base suggesting an association between inflammation and depression, and with a more limited evidence-base indicating an association between increased CRP levels mainly in subjects with generalized anxiety disorder (GAD), including our previous work from NESDA, ALSPAC, and UK Biobank cohorts [2, 3, 34–36, 4, 5, 9, 11, 17–19, 21]. Together, these findings support the idea that systemic inflammation could be specifically related to depressive rather than anxiety symptoms.

Our results provide evidence for further phenotype-specificity within the depression syndrome suggesting that inflammation maps specifically onto symptoms of fatigue, sleeping problems, changes in appetite, and depressed mood. These findings are consistent with previous research[10–15] including the concept of immuno-metabolic depression and with inflammation-related “sickness behaviour” observed in human and animal studies [37, 38].

We add to this evidence-base by providing data for similar and distinct associations for IL-6 and CRP for certain symptoms, especially sleep and appetite. Only hypersomnia, but not loss of sleep, showed consistent associations with CRP and IL-6. We report an intriguing dissociation between these inflammatory markers regarding their associations with appetite. CRP and IL-6 were specifically associated with increased and decreased appetite, respectively. These findings are consistent with evidence from animal models showing that CRP directly inhibits leptin binding to its central receptors, abolishing its anorexigenic effect and disinhibiting food intake [39]. In contrast, in obese mice with leptin resistance, the central activation of IL-6 trans-signalling has been shown to suppress feeding and improve glucose tolerance [40]. Similarly, it has been previously shown that increased BMI, a major stimulus for CRP production [41], is associated with appetite alterations [21], but only with increased appetite [42]. These data shed light on the impact of obesity on the association between CRP and appetite alterations, which was fully attenuated after controlling for BMI.

The pathways linking BMI, inflammation and depression, and their role in those pathways, are particularly complex. It is known that genetic risk variants for inflammation and for depression also have a major role in BMI increase [43, 44]; this may create a configuration in which BMI is both a confounder and a collider (Supplementary Figure3), whose adjustment or lack thereof may lead to biased estimates. Nevertheless, all associations with other symptoms remained statistically significant after BMI adjustment, although relatively reduced.

Findings from MR analyses also highlight divergence between inflammatory markers. IL-6 seems to have a potential causal effect in the development of fatigue and sleep problems, in line with evidence on the role of inflammation in “sickness behaviour” [37, 38]. In contrast, CRP did not show significant MR estimates for the same symptoms, despite strong associations with circulating protein levels. These results suggest that the CRP-depressive symptom associations may represent epiphenomena emerging from common underlying factors such as metabolic dysregulation. This idea is consistent with previous evidence suggesting shared genetic liability between CRP and symptoms of fatigue, altered sleep and appetite [15, 18, 21]. Furthermore, genetically-elevated CRP levels were associated with lower risk of psychological symptoms, cognitive problems and suicidality. Similar divergent MR results have also been reported for schizophrenia, suggesting a protective effect of CRP and a risk-increasing effect of soluble IL-6 receptor (IL-6R) on schizophrenia risk [45]. We have also previously reported that *IL-6R* variants associated with higher serum IL-6 levels (but decreased IL-6R activity) and *CRP* variants associated with higher CRP levels are *both* associated with increased risk of depression in the UK Biobank [46]. It could be hypothesized that the genetic instrument for CRP may partially capture the activity of IL-6 classic signalling that promotes regenerative and protective responses for neuronal function [47].

Hence, IL-6 classic signalling may potentially underlie protective findings for schizophrenia and depression. Conversely, IL-6 trans-signalling, which has been implicated in chronic inflammatory conditions such as rheumatoid arthritis and is primarily responsible for the pro-inflammatory role of IL-6 [48], may be responsible for risk-increasing associations of genetic instruments for IL-6. Future analyses joinlty considering CRP and IL-6 genetic instruments may clarify the direct effect of each marker. It needs to be noted that while MR analysis can provide evidence for causality, population genomic approaches alone are not sufficient to clarify pathophysiologic mechanisms fully. Triangulation of research evidence using different approaches including experimental studies of immune-modulation in humans and animals are required to fully elucidate potential role of the IL-6/IL-6R pathway and other immune biomarkers in the pathogenesis of these disorders. Notwithstanding these outstanding questions, our findings suggest that inflammation, and the IL-6/IL-6R pathway particularly, could be causally linked with risk of depression and schizophrenia.

### Strengths and Limitations

Strengths of the work include use of large samples from two European cohorts allowing replication/verification of findings, combined phenotypic and genetic analyses, and evaluation of both depressive and anxiety symptoms. A limitation was the lack of more granular data on specific vegetative depressive symptoms and IL-6 levels in UKB. Furthermore, as the overwhelming majority of subjects enrolled were of European ancestry results cannot be generalized to populations of other ancestries.

## CONCLUSIONS

In the current investigation of associations of inflammation with depression and anxiety, we provide evidence for specificity at several levels. First, systemic inflammation is mainly associated with depressive rather than anxiety symptoms. Second, within depression, inflammation is particularly associated with somatic/neurovegetative symptoms such as fatigue, altered sleep, appetite as well as depressed mood and anhedonia. Third, within symptoms, IL-6 and CRP have opposing effects on appetite but similar effects on fatigue. In addition, using MR analysis we provide evidence that the IL-6/IL-6R pathway could be causally linked with fatigue and sleeping problems. The field now requires experimental studies of IL-6 modulation in humans and animals to further evaluate causality, potential pathogenic mechanisms, and to assess potential usefulness of (add-on) immunotherapies for depression.

## Supporting information

STROBE checklist

## Data Availability

We provide data for genetic instruments and MR analysis scripts on the Open Science Framework under https://osf.io/2fwr6/ for full reproducibility.

## ACKNOWLEDGMENT

YM is partially supported by the Complex Trait Genetics programme of Amsterdam Neuroscience. NK and SM are supported by the International Max Planck Research School of Translational Psychiatry (IMPRS-TP). FL is supported by FP7–Marie Curie Career Integration Grant PCIG12-GA-2012-334065 from the European Union Seventh Framework Programme. GMK and PBJ acknowledge support from the MQ: Transforming Mental Health (Data Science Award; grant code: MQDS17/40), which also supported ZY. GMK also acknowledges funding support from the Wellcome Trust (grant code: 201486/Z/16/Z), the Medical Research Council UK (grant code: MC_PC_17213 and MR/S037675/1), and the BMA Foundation (J Moulton grant 2019). BWJHP has received research funding (unrelated to the work reported here) from Jansen Research and Boehringer Ingelheim. This research has been conducted using the UK Biobank resource.

## Notes

### Competing Interest Statement

The authors have declared no competing interest.

### Author Declarations

The UKB study was approved by the UK Biobank research ethics committee and Human Tissue Authority research tissue bank. The current analysis was approved under project no. 26999. The NESDA research protocol was approved centrally by the Vrije Universiteit (VU) Amsterdam University Medical Centre ethics committee and locally by the ethics committee of participating universities. Participants from both cohorts provided informed consent.

