## Supplementary Material 1 for "Association of Inflammation with Depression and Anxiety: Evidence for Symptom-Specificity and Potential Causality from UK Biobank and NESDA Cohorts"

ONLINE SUPPLEMENT

### SUPPLEMENTARY METHODS

#### Study cohorts

##### UK Biobank

The UK Biobank is a population-based cohort comprising 502,524 UK residents aged 40-69 years who were recruited between 2006 and 2010 from 22 assessment centres throughout the UK to reflect a broad socioeconomic demographic and mixture of urban and rural residents [1]. The full dataset includes a range of phenotyping assessments, biochemical assays and genome-wide genotyping of baseline samples from all participants. The UK Biobank study was approved by the UK Biobank’s research ethics committee and Human Tissue Authority research tissue bank. Informed consent was obtained from all participants.

In the present study, phenotypic analyses included up to 147,478 participants with data on CRP and depressive/anxiety symptoms (sample size varying per symptom) assessed as part of a follow-up mental health survey [2]. For Mendelian randomisation analysis, a sample of unrelated individuals with European ancestry, and genotype information was used. This resulted in inclusion of up to 325,441 participants to estimate exposure associations with CRP for one-sample MR analysis (see details below) and inclusion of up to 111,572 participants to estimate outcome associations.

##### NESDA

The Netherlands Study of Depression and Anxiety (NESDA) is an ongoing cohort study into the long-term course and consequences of depressive and anxiety disorders [3]. In 2004–2007, participants aged 18–65 years were recruited from the community (19%), general practice (54%) and secondary mental health care (27%). A total of 2,981 participants were included, consisting of persons with a current or past depressive and/or anxiety disorder and healthy control subjects, and were followed-up during biannual assessments. 2,905 participants with complete data at baseline on inflammatory markers and depressive/anxiety symptoms were selected. Biomarkers and symptoms were assessed again at 2-year and 6-year follow-up, totalling ~7,000 observations.

#### Depressive and anxiety symptoms

##### UK Biobank

In the UK Biobank study, symptoms were assessed using an online, follow-up mental health survey [2], which included the Patient Health Questionnaire-9 (PHQ-9)[4] and the General Anxiety Disorder-7 (GAD-7)[5] assessing, respectively, nine depressive and seven anxiety symptoms. Each item was scored on a 4-point scale, which was recoded/ binarised to reflect endorsement versus no endorsement of each symptom. For depressive symptoms we also created two summary scores, “psychological” and “somatic”, based on a two-factor model emerging from the genetic covariance between the nine symptoms [6].

##### NESDA

In NESDA, items investigating the corresponding symptoms assessed in UK Biobank were drawn from different questionnaires administered at baseline, 2- and 6-year follow-up: the Inventory of Depressive Symptomatology (IDS-SR_30_)[7], the Beck Anxiety Inventory (BAI)[8] and the Penn State Worry Questionnaire (PSWQ)[9].

The 30-item self-report IDS-SR_30_ measures specific depressive symptoms endorsed in the past week and their severity. Rating on 4-point Likert scales includes core symptoms of major depressive episodes, melancholic (e.g., anhedonia, nonreactive mood, psychomotor retardation/ agitation, appetite or weight decrease, early morning awakening, and self-outlook) and atypical (e.g., mood reactivity, leaden paralysis, weight gain or increased appetite, hypersomnia, and interpersonal sensitivity) features, and commonly associated symptoms (e.g., irritability, anxiety, somatic complaints).

The BAI is a short list describing 21 anxiety symptoms such as "wobbliness in legs", "scared" and "fear of losing control". Respondents are asked to rate how much each of these symptoms bothered them in the past week on a 4-point Likert scale, ranging from 1 (“not at all”) to 4 (“severely, I could barely stand it”).

The PSWQ is a 16-item self-report questionnaire that assesses the frequency and intensity of worry symptoms. Each item is rated on a 5-point Likert scale from 1 (“not at all typical”) to 5 (“very typical”).

##### Supplementary Table 1. Depressive and anxiety symptoms assessment

|  |  | **UK BIOBANK** | | |  | **NESDA** | | |
| --- | --- | --- | --- | --- | --- | --- | --- | --- |
| **Depressive Symptoms** |  | **Instrument** | **Item** | **Coding** |  | **Instrument** | **Item** | **Coding** |
| D.1 Anhedonia |  | PHQ-9 | 1. Recent lack of interest or pleasure in doing things | Range 0-3, ≥1 Endorsed |  | IDS-SR_30_ | Average items (19,21) | Range 1-4, ≥3 Endorsed |
|  |  |  |  |  |  | IDS-SR_30_ | 19. General Interest | Range 1-4, ≥3 Endorsed |
|  |  |  |  |  |  | IDS-SR_30_ | 21. Capacity for Pleasure or Enjoyment (excluding sex) | Range 1-4, ≥3 Endorsed |
| D.2 Depressed mood |  | PHQ-9 | 2. Recent feelings of depression | Range 0-3, ≥1 Endorsed |  | IDS-SR_30_ | 5. Feeling Sad | Range 1-4, ≥3 Endorsed |
| D.3 Sleep problems |  | PHQ-9 | 3. Trouble sleeping, or sleeping too much | Range 0-3, ≥1 Endorsed |  | IDS-SR_30_ | Average items (1,2,3,4) | Range 1-4, ≥3 Endorsed |
|  |  |  |  |  |  | IDS-SR_30_ | 1. Falling Asleep | Range 1-4, ≥3 Endorsed |
|  |  |  |  |  |  | IDS-SR_30_ | 2. Sleep During the Night | Range 1-4, ≥3 Endorsed |
|  |  |  |  |  |  | IDS-SR_30_ | 3. Waking Up Too Early | Range 1-4, ≥3 Endorsed |
|  |  |  |  |  |  | IDS-SR_30_ | 4. Sleeping Too Much | Range 1-4, ≥3 Endorsed |
| D.4 Fatigue |  | PHQ-9 | 4. Recent feelings of tiredness or low energy | Range 0-3, ≥1 Endorsed |  | IDS-SR_30_ | Average items (20,30) | Range 1-4, ≥3 Endorsed |
|  |  |  |  |  |  | IDS-SR_30_ | 20. Energy Level | Range 1-4, ≥3 Endorsed |
|  |  |  |  |  |  | IDS-SR_30_ | 30. Leaden Paralysis | Range 1-4, ≥3 Endorsed |
| D.5 Appetite changes |  | PHQ-9 | 5. Recent poor appetite or overeating | Range 0-3, ≥1 Endorsed |  | IDS-SR_30_ | Average items (11,12) | Range 1-4, ≥3 Endorsed |
|  |  |  |  |  |  | IDS-SR_30_ | 11. Decreased Appetite | Range 1-4, ≥3 Endorsed |
|  |  |  |  |  |  | IDS-SR_30_ | 12. Increased Appetite | Range 1-4, ≥3 Endorsed |
| D.6 Feelings of inadequacy |  | PHQ-9 | 6. Recent feelings of inadequacy | Range 0-3, ≥1 Endorsed |  | IDS-SR_30_ | 16. View of Myself | Range 1-4, ≥3 Endorsed |
| D.7 Cognitive problems |  | PHQ-9 | 7. Recent trouble concentration | Range 0-3, ≥1 Endorsed |  | IDS-SR_30_ | 15. Concentration/Decision Making | Range 1-4, ≥3 Endorsed |
| D.8 Psychomotor changes |  | PHQ-9 | 8. Recent changes in moving/speaking | Range 0-3, ≥1 Endorsed |  | IDS-SR_30_ | Average items (23,24) | Range 1-4, ≥3 Endorsed |
|  |  |  |  |  |  | IDS-SR_30_ | 23. Feeling slowed down | Range 1-4, ≥3 Endorsed |
|  |  |  |  |  |  | IDS-SR_30_ | 24. Feeling restless* | Range 1-4, ≥3 Endorsed |
| D.9 Suicidal ideation |  | PHQ-9 | 9. Recent thoughts of suicide or self-harm | Range 0-3, ≥1 Endorsed |  | IDS-SR_30_ | 18. Thoughts of Death or Suicide | Range 1-4, ≥3 Endorsed |
| **Anxiety Symptoms** |  | **Instrument** | **Item** | **Coding** |  | **Instrument** | **Item** | **Coding** |
| A.1 Anxiety |  | GAD-7 | 1. Recent feelings or nervousness or anxiety | Range 0-3, ≥1 Endorsed |  | IDS-SR_30_ | 7. Feeling Anxious or Tense: | Range 1-4, ≥3 Endorsed |
| A.2 Worrying control |  | GAD-7 | 2. Recent inability to stop or control worrying | Range 0-3, ≥1 Endorsed |  | PSWQ | Q9. Once I start worrying, I cannot stop. | Range 1-5, ≥4 Endorsed |
| A.3 Generalized worrying |  | GAD-7 | 3. Recent worrying too much about different things | Range 0-3, ≥1 Endorsed |  | PSWQ | Q2. Many situations make me worry. | Range 1-5, ≥4 Endorsed |
| A.4 Lack of relaxation |  | GAD-7 | 4. Recent trouble relaxing | Range 0-3, ≥1 Endorsed |  | BAI | Q4. Unable to relax | Range 1-4, ≥3 Endorsed |
| A.5 Restlessness |  | GAD-7 | 5. Recent restlessness | Range 0-3, ≥1 Endorsed |  | IDS-SR_30_ | 24. Feeling restless* | Range 1-4, ≥3 Endorsed |
| A.6 Irritability |  | GAD-7 | 6. Recent easy annoyance or irritability | Range 0-3, ≥1 Endorsed |  | IDS-SR_30_ | 6. Feeling Irritable | Range 1-4, ≥3 Endorsed |
| A.7 Foreboding |  | GAD-7 | 7. Recent feelings of foreboding | Range 0-3, ≥1 Endorsed |  | BAI | Q5. Fear of worst happening | Range 1-4, ≥3 Endorsed |
| **Summary symptom scores** | | | | | | | |  |
| **DEPRESSION Psychological** = D.1 + D.2 + D.6 + D.9 | | | | | | | |  |
| **DEPRESSION Somatic** = D.3 + D.4 + D.5 + D.7 + D.8 | | | | | | | |  |

*Note*: *The item **“**Feeling restless**”** from IDS-SR_30_ is used in both D.8 Psychomotor changes and A.5 Restlessness

#### Inflammatory markers

##### UK Biobank

Serum high-sensitivity C-reactive protein (CRP) levels were measured as part of a large biomarker assay effort for the UK Biobank that included baseline serum samples from all 500,000 individuals as well as 20,000 individuals who attended a repeat follow-up assessment. High-sensitivity CRP was measured using the Immuno-turbidimetric system from the Beckman Coulter AU5800 analytical platform (Beckman Coulter Inc., Brea, CA, USA). The manufacturer’s analytical range was 0.08-80mg/L.

Further information is available from official UK Biobank documentation on CRP specifically (<https://biobank.ndph.ox.ac.uk/showcase/field.cgi?id=30710>) and the companion document for biomarker data (<https://biobank.ndph.ox.ac.uk/showcase/refer.cgi?id=1227>).

##### NESDA

Inflammatory markers were determined from fasting morning blood plasma at baseline, 2- and 6-year follow-up. Plasma levels of CRP at baseline were measured in duplicate by an inhouse high-sensitivity enzyme-linked immunosorbent assay (ELISA) based on purified protein and polyclonal anti-CRP antibodies (Dako, Glostrup, Denmark). The lower detection limit of CRP is 0.1 mg/L and the sensitivity is 0.05 mg/ L. Intra- and inter-assay coefficients of variation were 5% and 10%, respectively. Plasma levels of CRP in the follow-up waves were measured in duplicate by a high-sensitivity particle enhanced immunoturbidimetric assay (CRPHS, Roche Diagnostics, Indianapolis, IN, USA). The lower detection limit of CRP in this kit is 0.15 mg/L and the sensitivity is 0.3 mg/L. Intra- and inter-assay coefficients of variation were as follows. Intra-assay: 2-yr FU 5%; 6-yr FU 7%; Inter-assay 2-yr FU 4%; 6-yr FU 9%. Plasma IL-6 levels at baseline were measured in duplicate by a high-sensitivity ELISA (PeliKine CompactTM ELISA, Sanquin, Amsterdam, the Netherlands). The lower detection limit of IL- 6 is 0.35 pg/ml and the sensitivity is 0.10 pg/ml. Intra- and inter-assay coefficients of variation were 8% and 12%, respectively. At the 2- and 6-year follow-up, IL-6 was measured in duplicate by a high-sensitivity solid-phase ELISA (Human IL-6 Quantikine HS kit, R&D Systems, Minneapolis, MN, USA). The lower detection limit of IL-6 in this kit is 0.08 pg/ml and the sensitivity range is 0.016–0.110 pg/ml. Intra- and inter-assay coefficients of variation were 7.8% and 7.2%, respectively.

#### Covariates

##### UK Biobank

In the UK Biobank, covariates were measured during the baseline assessment. In addition to sociodemographic characteristics of age and sex, sociodemographic status (SES) was assessed using Townsend Deprivation Index (TDI) as proxy phenotype. The Townsend Deprivation Index constitutes a measure of deprivation based on an individual’s postcode that is referenced against census data and incorporates information on employment, house and car ownership, and household crowing [10]. Smoking was self-reported as “never”, “previous”, “current”, or “prefer not to answer”. Alcohol use was self-reported as “daily or almost daily”, “three or four times a week”, “once or twice a week”, “one to three times a month”, “special occasions only”, “never”, or “prefer not to answer”. Physical activity was self-reported as type of activity during the past 4 weeks including “walking for pleasure (not as a means of transport)”, “other exercises (e.g.: swimming, cycling, keep fit, bowling)”, “strenuous sports”, “light DIY (e.g.: pruning, watering the lawn)”, “heavy DIY (e.g.: weeding, lawn mowing, carpentry, digging)”, “none of the above”, or “prefer not to answer”. History of diabetes was self-reported as diagnosed by a doctor and coded as “yes”, “no”, “do not know”, and “prefer not to say”. History of cardiovascular disease was self-reported for conditions of “heart attack”, “angina”, “stroke”, “high blood pressure”, “none of the above”, or “prefer not to answer” as diagnosed by a doctor. Body Mass Index (BMI) was coded using the standard formula kg/m².

Further information is available online from relevant UK Biobank data fields per covariate; sex (data field [31](https://biobank.ctsu.ox.ac.uk/crystal/field.cgi?id=31)), age (data field [21003](https://biobank.ctsu.ox.ac.uk/crystal/field.cgi?id=21003)), Townsend Deprivation Index (data field [189](https://biobank.ctsu.ox.ac.uk/crystal/field.cgi?id=189)), smoking (data field [20116](https://biobank.ctsu.ox.ac.uk/crystal/field.cgi?id=20116)), alcohol use (data field [1558](https://biobank.ctsu.ox.ac.uk/crystal/field.cgi?id=1558)), physical activity (data field [6164](https://biobank.ctsu.ox.ac.uk/crystal/field.cgi?id=6164)), diabetes (data field [2443](https://biobank.ctsu.ox.ac.uk/crystal/field.cgi?id=2443)), cardiovascular disease (data field [6150](https://biobank.ctsu.ox.ac.uk/crystal/field.cgi?id=6150)), and BMI (data field [21001](https://biobank.ctsu.ox.ac.uk/crystal/field.cgi?id=21001)).

##### Supplementary Table 2. Covariate distributions in UK Biobank

|  |  | **UK Biobank** |  |
| --- | --- | --- | --- |
| **Characteristics** |  | ***N = 143,465*** | ***Missing, %*** |
| *Sociodemographics* |  |  |  |
| **Age** *years (mean ± SD)* |  | 55.87 (7.73) | 0 |
| **Sex (F)** *(%)* |  | 56.2 | 0 |
| **Townsend Deprivation Index** *(mean ± SD)* |  | -1.72 (2.83) | 0.1 |
| *Lifestyles & Health* |  |  |  |
| **Smoking status** *(%)* |  |  | 0 |
| prefer not to answer |  | 0.2 |  |
| never |  | 57.4 |  |
| former |  | 35.2 |  |
| current |  | 7.2 |  |
| **Alcohol consumption** *(%)* |  |  | 0.1 |
| never |  | 5.5 |  |
| occasional |  | 45.0 |  |
| regular |  | 49.6 |  |
| **Physical activity** *(%)* |  |  | 4.0 |
| Walking for pleasure |  | 78.9 |  |
| Other exercises |  | 12.6 |  |
| Strenuous sports |  | 0.9 |  |
| Light DIY |  | 5.8 |  |
| Heavy DIY |  | 1.8 |  |
| **T2D/CVD** *(%)* |  | 24.8 | 0 |
| **BMI** *(kg/m^2^) (mean ± SD)* |  | 26.77 (4.54) | 0.2 |

*Note*: DIY=Do-it-yourself

##### NESDA

Covariates for the present study were measured at baseline. Sociodemographic characteristics included age, sex, and years of education. The following lifestyle characteristics were assessed: being a current smoker; alcohol consumption measured as drinks/week and categorized as non-drinker, mild-moderate drinker, (1–14 women/1–21 men), and heavy drinker (>14 women / >21 men); physical activity measured with the International Physical Activity Questionnaire[11] and expressed in metabolic equivalent minutes (ratio of energy expenditure during activity compared with rest times the number of minutes performing the activity) per week. History of diabetes, vascular and heart problems was assessed via self-report. Height and weight were measured to calculate body mass index (BMI) in kg/m^2^.

##### Supplementary Table 3. Covariate distributions in NESDA

|  |  | **NESDA** |
| --- | --- | --- |
| **Characteristics** |  | ***N = 2,905*** |
| *Sociodemographics* |  |  |
| **Age** *years (mean ± SD)* |  | 41.9 (13.1) |
| **Sex (F)** *(%)* |  | 66.5 |
| **Education** *years (mean ± SD)* |  | 12.2 (3.3) |
| *Lifestyles & Health* |  |  |
| **Smoking status** *(%)* |  |  |
| never |  | 28.0 |
| former |  | 33.6 |
| current |  | 38.4 |
| **Alcohol consumption** *(%)* |  |  |
| non-drinker |  | 32.3 |
| mild-moderate |  | 56.1 |
| heavy |  | 11.6 |
| **Physical activity** *METmin/week/1000 (mean ± SD)* |  | 3.7 (3.0) |
| **T2D/CVD** *(%)* |  | 7.5 |
| **BMI** *(kg/m^2^) (mean ± SD)* |  | 25.6 (5.0) |

#### Phenotypic association analyses

In both cohorts the association between inflammatory marker levels and depressive/anxiety symptoms was estimated by regressing individual symptoms and summary scales on log-transformed values of the markers. UK Biobank analyses used logistic and linear regression models for, respectively, individual symptoms and summary scales. In NESDA, generalized linear mixed models (for individual symptoms) and linear mixed models (for summary scales) were used with random intercept at the subject level to take into account the correlation between repeated measures within subjects. All models in NESDA were adjusted for wave of assessment.

Analyses in UK Biobank were conducted using Stata/SE 16.0 (Stata, College Station, TX) or R v3.6.1 (R Project for Statistical Computing)[12].

Association analyses in NESDA were performed in *R* v4.0.0 (R Project for Statistical Computing)[12] using the *lme4* (v1.1-23) package [13].

Example of generalized linear mixed models for individual symptom:

assoc <- glmer(symptom ~ (log)CRP + assessment_wave + covariates + (1 | ID), data = dataset, family = binomial, control = glmerControl(optimizer = "bobyqa"), nAGQ = 10)

Example of linear mixed models for summary scales:

assoc <- lmer(symptom ~ (log)CRP + assessment_wave + covariates + (1 | ID), data = dataset)

Cohort-specific estimates were pooled in *R* (v4.0.3) and the *metafor* (v2.4-0) package[14] using random-effects meta-analysis with the DerSimonian and Laird method [15]. Example of estimate meta-analysis:

rma_fit <- rma(yi = est, sei = se, data = dataset, method = "DL")

All statistical tests were two-sided and used a significance level of *P*<0.05. False-Discovery Rate (FDR) q-values were calculated taking into account multiple testing as described in the main text using the ‘p.adjust’ function in *R*.

#### Mendelian randomisation analysis

##### Genotyping and quality control

For MR analyses, the imputed UK Biobank genetic data (version 3) for ~488,000 individuals were used. Samples were genotyped on the UK BiLEVE Axiom Array (~45,000 samples) or the Affymetrix UK Biobank Axiom Array (~443,000 samples). Quality control (QC) before imputation was performed centrally followed by imputation as described by Bycroft *and colleagues* [16]. For post-imputation QC, we excluded individuals with non-British ancestry and individuals related up to the third degree resulting in a final dataset of 342,081 individuals. Per-Variant QC was performed using QCTOOL (<https://enkre.net/cgi-bin/code/qctool/dir?ci=trunk>) to exclude SNPs with minor allele frequency (MAF) < 1% , imputation score < 60%, genotyping missingness > 2%, and Hardy-Weinberg Equilibrium (HWE) test p-value < 0.000001.

##### Instrument selection

We defined multiple genetic proxy variables for CRP and IL-6 based on prior reports.[17–20] As proxy instruments for CRP, we selected 24 SNPs[17] and 4 SNPs[20] located in the CRP gene and associated with CRP levels. As proxy instruments for IL-6, we selected SNPs from the IL-6R gene; 7 SNPs[17] associated with CRP levels as a downstream readout of IL-6 signalling as well as 3 SNPs[19] and 1 SNP[18] associated with IL-6 levels. Number of SNPs and associated sample sizes for 1-sample and 2-sample MR analyses are provided in Supplementary Table 4.

##### Mendelian randomisation analysis

MR analysis was conducted in *R* (v4.0.3) using the *TwoSampleMR* (v0.5.5) package [12, 21].

Availability of CRP concentrations in the UK Biobank sample allowed a 2-fold validation of MR analyses using both a two-sample and one-sample approach that regressed instrument SNPs on CRP and CRP/IL-6 depending on the instrument.

For the two-sample approach, GWAS effect estimates of SNPs on the exposure were taken from original GWAS reports[17–20] assessing genetic associations with CRP and IL-6 levels. For the one-sample approach, SNP effect estimates were calculated by regressing (log)CRP levels on the same individual SNPs used in two-sample MR while adjusting for age, age^2^, sex, age*sex, and 20 genotype principal components. For both approaches, effect estimates of SNPs on outcome phenotypes were calculated using the same linear regression approach. Variant harmonisation for the two-sample approach was conducted by excluding ambiguous and palindromic SNPs, for which strand information could not be inferred [22].

Next, fixed-effects inverse variance weighted (IVW) MR analysis was conducted per exposure-outcome combination for all instruments except the Sarwar IL-6 instrument [18], which was only based on a single SNP (rs2228145); here, a Wald ratio estimate was computed [23, 24]. Cochrane’s *Q* was estimated to provide an indication on potential horizontal pleiotropy [25].

##### Availability of code and genetic instrument data

We provide data for genetic instruments and MR analysis scripts on the Open Science Framework under <https://osf.io/2fwr6/> for full reproducibility.

##### Supplementary Table 4. Genetic instruments for MR analysis

|  | **2-Sample MR** | | | | **1-Sample MR** | | | |
| --- | --- | --- | --- | --- | --- | --- | --- | --- |
| **Instrument** | **Phenotype** | **Number of SNPs** | **Instrument Strength (F-statistic)^a^** | **Sample Size^b^** | **Phenotype** | **Number of SNPs** | **Instrument Strength (F-statistic)^a^** | **Sample Size^b^** |
| ***CRP*** |  |  |  |  |  |  |  |  |
| Georgakis *et al.*[17] | CRP | 24 | Min=55; Median=274; Max=3413 | 204.402 | CRP | 24 | Min=24; Median=160; Max=4573 | 304,610-325,441 |
| CCGC[20] | CRP | 3 | Min=236; Median=649; Max=1349 | 38,573-105,476 | CRP | 4 | Min=1750; Median=2510; Max=4573 | 322,826-325,441 |
| ***IL-6*** |  |  |  |  |  |  |  |  |
| Georgakis *et al.*[17] | CRP | 7 | Min=80; Median=138; Max=764 | 204.402 | CRP | 7 | Min=127; Median=175; Max=1509 | 299,591-325,441 |
| Swerdlow *et al.*[19] | IL-6 | 3 | Min=8;  Median=14;  Max=16 | 4,462-4,479 | CRP | 3 | Min=618; Median=620; Max=1489 | 323,494-325,369 |
| Sarwar *et al.*[18] | IL-6 | 1 | 397^c^ | 27.185 | CRP | 1 | 1510 | 325,441 |

*Note*: ^a^Instrument strength F-statistics are based on the formulae $R^{2}=2*MAF*\left( 1-MAF \right)*{beta}^{2}$, where MAF=Minor allele frequency, and $F= \frac{R^{2}* ( N-2 )}{1-R^{2}}$ ; as described in Shim *et al.*[26] and Palmer *et al.*[27] previously. ^b^Sample sizes are provided as range as this can be varying per SNP. If single values are presented, there is no variability. ^c^Since MAF/ effect allele frequency were not available, the approximate F-statistic was calculated as $F= \frac{{beta}^{2}}{{se}^{2}}$ as described in Rosa *et al.*[28] and Pierce *et al.*[29] previously.

### SUPPLEMENTARY RESULTS

##### Supplementary Table 5. Main variables at 2- and 6-year follow-up in NESDA

|  |  | **NESDA** | | |
| --- | --- | --- | --- | --- |
|  |  | **2-year follow-up** |  | **6-year follow-up** |
| *Symptoms endorsement - Depression* (N,%) |  |  |  |  |
| **D.1 Anhedonia** |  | 169 (6.89%) |  | 141 (6.70%) |
| **D.2 Depressed mood** |  | 288 (11.8%) |  | 220 (10.4%) |
| **D.3 Sleep problem** |  | 436 (17.8%) |  | 406 (19.3%) |
| **D.4 Fatigues** |  | 533 (21.7%) |  | 423 (20.1%) |
| **D.5 Appetite change** |  | 223 (7.71%) |  | 72 (3.43%) |
| **D.6 Feelings of inadequacy** |  | 358 (14.7%) |  | 284 (13.5%) |
| **D.7 Cognitive problems** |  | 283 (11.6%) |  | 218 (10.4%) |
| **D.8 Psychomotor change** |  | 222 (9.05%) |  | 184 (8.73%) |
| **D.9 Suicidal ideation** |  | 171 (6.99%) |  | 284 (13.5%) |
| **Psychological symptom score** *(mean, SD)* |  | 0.402 (0.814) |  | 0.376 (0.790) |
| **Somatic symptom score** *(mean, SD)* |  | 0.643 (0.997) |  | 0.617 (1.01) |
| *Symptoms endorsement - Anxiety* (N,%) |  |  |  |  |
| **A.1 Anxiety** |  | 321 (13.1%) |  | 238 (11.3%) |
| **A.2 Worrying control** |  | 493 (21.8%) |  | 388 (18.5%) |
| **A.3 Generalized worrying** |  | 633 (27.8%) |  | 454 (21.6%) |
| **A.4 Lack of relaxation** |  | 486 (19.8%) |  | 390 (18.6%) |
| **A.5 Restlessness** |  | 418 (17.1%) |  | 418 (17.1%) |
| **A.6 Irritability** |  | 275 (11.2%) |  | 221 (10.5%) |
| **A.7 Foreboding** |  | 277 (11.3%) |  | 177 (8.42%) |
| *Inflammatory markers* |  |  |  |  |
| **CRP (mg/L)** *(median, IQR)* |  | 1.07 (0.43 - 2.42) |  | 1.14 (0.52 - 2.71) |
| **IL-6 (pg/mL)** *(median, IQR)* |  | 1.02 (0.67 - 1.67) |  | 0.94 (0.59 - 1.57) |

##### Supplementary Figure 1. Repeated measures of CRP across NESDA assessment waves

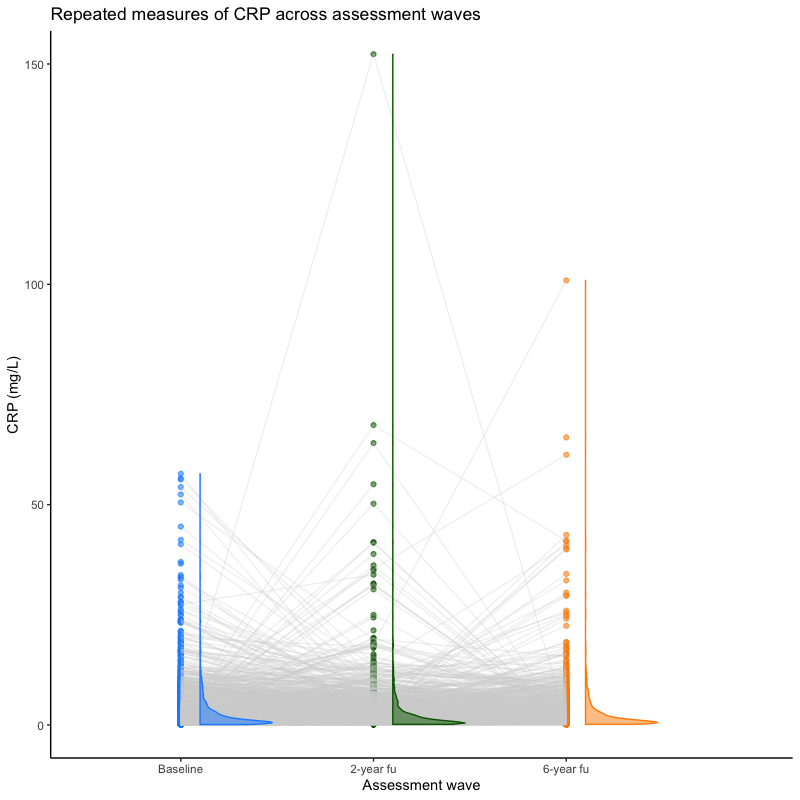

##### Supplementary Figure 2. Repeated measures of IL-6 across NESDA assessment waves

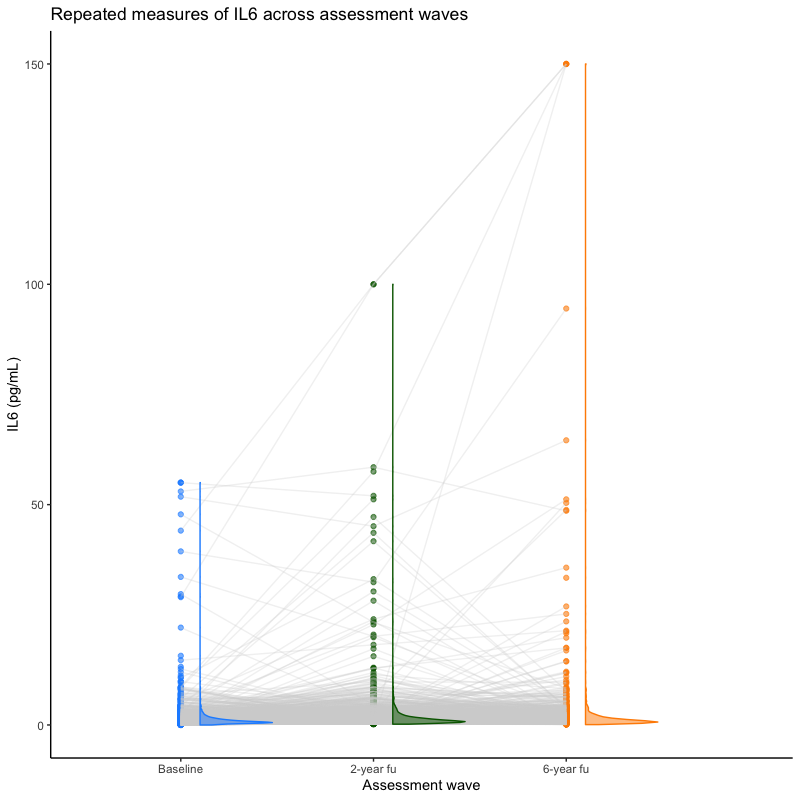

#### Cross-cohort analyses on the association between CRP and depressive/anxiety symptoms

##### Supplementary Table 6. UK Biobank Association Results between CRP and Depressive/Anxiety Symptoms

|  |  | **Model 1** |  |  | **Model 2** |  |  | **Model 3** |  |
| --- | --- | --- | --- | --- | --- | --- | --- | --- | --- |
| **Symptom** | **OR (95% CI)** | **P** | **Q** | **OR (95% CI)** | **P** | **Q** | **OR (95% CI)** | **P** | **Q** |
| ***Depressive symptoms*** |  |  |  |  |  |  |  |  |  |
| Anhedonia | 1.16 (1.15-1.18) | <0.001 | <0.001 | 1.11 (1.1-1.13) | <0.001 | <0.001 | 1.05 (1.04-1.07) | <0.001 | <0.001 |
| Depressed mood | 1.1 (1.09-1.12) | <0.001 | <0.001 | 1.07 (1.05-1.08) | <0.001 | <0.001 | 1.03 (1.01-1.04) | <0.001 | <0.001 |
| Sleeping problems | 1.07 (1.06-1.08) | <0.001 | <0.001 | 1.05 (1.04-1.06) | <0.001 | <0.001 | 1.02 (1.01-1.03) | 0.001 | 0.002 |
| Fatigue | 1.17 (1.16-1.18) | <0.001 | <0.001 | 1.12 (1.11-1.13) | <0.001 | <0.001 | 1.06 (1.05-1.07) | <0.001 | <0.001 |
| Appetite changes | 1.33 (1.31-1.34) | <0.001 | <0.001 | 1.26 (1.24-1.27) | <0.001 | <0.001 | 1.04 (1.03-1.06) | <0.001 | <0.001 |
| Feelings of inadequacy | 1.07 (1.06-1.08) | <0.001 | <0.001 | 1.04 (1.03-1.05) | <0.001 | <0.001 | 1 (0.99-1.02) | 0.733 | 0.822 |
| Cognitive problems | 1.11 (1.09-1.12) | <0.001 | <0.001 | 1.07 (1.05-1.08) | <0.001 | <0.001 | 1.02 (1-1.03) | 0.033 | 0.054 |
| Psychomotor changes | 1.17 (1.15-1.2) | <0.001 | <0.001 | 1.11 (1.09-1.14) | <0.001 | <0.001 | 1.05 (1.02-1.08) | <0.001 | <0.001 |
| Suicidal ideation | 1.14 (1.11-1.17) | <0.001 | <0.001 | 1.07 (1.05-1.1) | <0.001 | <0.001 | 1.03 (1-1.06) | 0.084 | 0.126 |
| ***Anxiety symptoms*** |  |  |  |  |  |  |  |  |  |
| Anxiety | 1.01 (1-1.03) | 0.013 | 0.013 | 1 (0.99-1.01) | 0.761 | 0.761 | 1 (0.99-1.02) | 0.565 | 0.678 |
| Worrying control | 1.06 (1.04-1.07) | <0.001 | <0.001 | 1.03 (1.02-1.04) | <0.001 | <0.001 | 1.02 (1-1.03) | 0.024 | 0.043 |
| Generalised worrying | 1.03 (1.02-1.04) | <0.001 | <0.001 | 1.01 (1-1.02) | 0.266 | 0.282 | 1 (0.99-1.01) | 0.776 | 0.822 |
| Lack of relaxation | 1.04 (1.02-1.05) | <0.001 | <0.001 | 1.01 (1-1.02) | 0.043 | 0.048 | 1.01 (1-1.02) | 0.165 | 0.212 |
| Restlessness | 1.06 (1.04-1.08) | <0.001 | <0.001 | 1.03 (1.01-1.04) | 0.002 | 0.002 | 1.01 (1-1.03) | 0.12 | 0.166 |
| Irritability | 1.09 (1.07-1.1) | <0.001 | <0.001 | 1.06 (1.05-1.07) | <0.001 | <0.001 | 1.03 (1.01-1.04) | <0.001 | <0.001 |
| Foreboding | 1.06 (1.05-1.08) | <0.001 | <0.001 | 1.03 (1.02-1.04) | <0.001 | <0.001 | 1 (0.99-1.02) | 0.874 | 0.874 |
| ***Depressive symptom score*** | **β (SE)** | **P** | **Q** | **β (SE)** | **P** | **Q** | **β (SE)** | **P** | **Q** |
| Psychological symptoms | 0.085 (0.004) | <0.001 | <0.001 | 0.051 (0.004) | <0.001 | <0.001 | 0.017 (0.004) | <0.001 | <0.001 |
| Somatic symptoms | 0.249 (0.006) | <0.001 | <0.001 | 0.176 (0.006) | <0.001 | <0.001 | 0.068 (0.007) | <0.001 | <0.001 |

##### Supplementary Table 7. NESDA Association Results between CRP and Depressive/Anxiety Symptoms

|  |  | **Model 1** |  |  | **Model 2** |  |  | **Model 3** |  |
| --- | --- | --- | --- | --- | --- | --- | --- | --- | --- |
| **Symptom** | **OR (95% CI)** | **P** | **Q** | **OR (95% CI)** | **P** | **Q** | **OR (95% CI)** | **P** | **Q** |
| ***Depressive symptoms*** |  |  |  |  |  |  |  |  |  |
| Anhedonia | 1.06 (0.95-1.18) | 0.329 | 0.586 | 1.01 (0.91-1.13) | 0.852 | 0.965 | 0.97 (0.86-1.09) | 0.575 | 0.762 |
| Depressed mood | 1.05 (0.96-1.15) | 0.264 | 0.572 | 1.02 (0.93-1.11) | 0.672 | 0.864 | 1.01 (0.93-1.11) | 0.762 | 0.762 |
| Sleeping problems | 1.08 (1-1.17) | 0.042 | 0.125 | 1.06 (0.99-1.15) | 0.106 | 0.278 | 1.06 (0.98-1.14) | 0.168 | 0.514 |
| Fatigue | 1.17 (1.08-1.26) | <0.001 | 0.002 | 1.12 (1.04-1.21) | 0.004 | 0.036 | 1.09 (1.01-1.18) | 0.037 | 0.337 |
| Appetite changes | 1.22 (1.08-1.37) | <0.001 | 0.009 | 1.18 (1.06-1.33) | 0.004 | 0.036 | 0.96 (0.85-1.08) | 0.514 | 0.762 |
| Feelings of inadequacy | 0.96 (0.88-1.04) | 0.286 | 0.572 | 0.94 (0.86-1.02) | 0.128 | 0.288 | 0.93 (0.86-1.02) | 0.132 | 0.514 |
| Cognitive problems | 1.02 (0.93-1.13) | 0.617 | 0.854 | 1 (0.91-1.1) | 0.965 | 0.967 | 0.98 (0.89-1.08) | 0.73 | 0.762 |
| Psychomotor changes | 0.96 (0.87-1.05) | 0.358 | 0.586 | 0.93 (0.85-1.03) | 0.155 | 0.31 | 0.91 (0.83-1.01) | 0.079 | 0.475 |
| Suicidal ideation | 0.94 (0.84-1.04) | 0.226 | 0.572 | 0.92 (0.83-1.02) | 0.108 | 0.278 | 0.93 (0.83-1.04) | 0.2 | 0.514 |
| ***Anxiety symptoms*** |  |  |  |  |  |  |  |  |  |
| Anxiety | 1.03 (0.95-1.11) | 0.503 | 0.755 | 1 (0.92-1.08) | 0.967 | 0.967 | 0.99 (0.9-1.07) | 0.73 | 0.762 |
| Worrying control | 1.01 (0.93-1.1) | 0.813 | 0.952 | 0.99 (0.91-1.08) | 0.858 | 0.965 | 0.98 (0.9-1.07) | 0.637 | 0.762 |
| Generalised worrying | 0.99 (0.92-1.08) | 0.899 | 0.952 | 0.98 (0.9-1.06) | 0.565 | 0.848 | 0.98 (0.9-1.07) | 0.625 | 0.762 |
| Lack of relaxation | 1.01 (0.94-1.09) | 0.766 | 0.952 | 0.98 (0.91-1.06) | 0.651 | 0.864 | 0.98 (0.9-1.06) | 0.562 | 0.762 |
| Restlessness | 0.9 (0.84-0.98) | 0.01 | 0.045 | 0.9 (0.83-0.97) | 0.006 | 0.036 | 0.9 (0.83-0.98) | 0.014 | 0.247 |
| Irritability | 1.11 (1.02-1.21) | 0.019 | 0.068 | 1.09 (1-1.18) | 0.06 | 0.216 | 1.07 (0.97-1.17) | 0.174 | 0.514 |
| Foreboding | 0.99 (0.91-1.08) | 0.863 | 0.952 | 0.97 (0.89-1.05) | 0.428 | 0.771 | 0.95 (0.88-1.04) | 0.286 | 0.573 |
| ***Depressive symptom score*** | **β (SE)** | **P** | **Q** | **β (SE)** | **P** | **Q** | **β (SE)** | **P** | **Q** |
| Psychological symptoms | 0.000 (0.009) | 0.992 | 0.992 | -0.006 (0.009) | 0.487 | 0.797 | -0.007 (0.009) | 0.423 | 0.762 |
| Somatic symptoms | 0.030 (0.011) | 0.005 | 0.029 | 0.022 (0.01) | 0.04 | 0.18 | 0.012 (0.011) | 0.26 | 0.573 |

##### Supplementary Table 8. Extended NESDA Association Results between CRP and Depressive/Anxiety Symptoms

|  |  | **Model 1** |  |  | **Model 2** |  |  | **Model 3** |  |
| --- | --- | --- | --- | --- | --- | --- | --- | --- | --- |
| **Symptom** | **OR (95% CI)** | **P** | **Q** | **OR (95% CI)** | **P** | **Q** | **OR (95% CI)** | **P** | **Q** |
| ***Depressive symptoms*** |  |  |  |  |  |  |  |  |  |
| Anhedonia |  |  | 0.659 |  |  | 0.967 |  |  | 0.824 |
| Depressed mood |  |  | 0.629 |  |  | 0.967 |  |  | 0.838 |
| *Sleeping problems* |  |  |  |  |  |  |  |  |  |
| Falling Asleep | 1.03 (0.95-1.13) | 0.437 | 0.791 | 1.01 (0.92-1.09) | 0.897 | 0.967 | 1 (0.92-1.09) | 0.962 | 0.962 |
| During the Night | 1.05 (0.99-1.11) | 0.092 | 0.288 | 1.05 (0.99-1.11) | 0.08 | 0.252 | 1.05 (0.99-1.11) | 0.14 | 0.573 |
| Waking Up Too Early | 0.99 (0.91-1.08) | 0.886 | 0.942 | 0.99 (0.91-1.07) | 0.76 | 0.967 | 1.01 (0.92-1.1) | 0.91 | 0.953 |
| Sleeping Too Much | 1.32 (1.17-1.48) | <0.001 | <0.001 | 1.27 (1.13-1.43) | <0.001 | 0.002 | 1.26 (1.11-1.43) | <0.001 | 0.006 |
| Fatigue |  |  | 0.001 |  |  | 0.031 |  |  | 0.274 |
| *Appetite changes* |  |  |  |  |  |  |  |  |  |
| Decreased | 1.05 (0.91-1.22) | 0.5 | 0.791 | 1.01 (0.87-1.17) | 0.915 | 0.967 | 1.1 (0.94-1.28) | 0.222 | 0.573 |
| Increased | 1.23 (1.1-1.37) | <0.001 | 0.003 | 1.21 (1.08-1.35) | 0.001 | 0.013 | 0.96 (0.85-1.09) | 0.556 | 0.824 |
| Feelings of inadequacy |  |  | 0.629 |  |  | 0.313 |  |  | 0.573 |
| Cognitive problems |  |  | 0.905 |  |  | 0.967 |  |  | 0.838 |
| *Psychomotor changes* |  |  |  |  |  |  |  |  |  |
| Feeling slowed down | 1.01 (0.92-1.11) | 0.84 | 0.942 | 0.99 (0.9-1.08) | 0.765 | 0.967 | 0.95 (0.86-1.04) | 0.261 | 0.573 |
| Feeling restless | 0.9 (0.84-0.98) | 0.01 | 0.044 | 0.9 (0.83-0.97) | 0.006 | 0.033 | 0.9 (0.83-0.98) | 0.014 | 0.151 |
| Suicidal ideation |  |  | 0.62 |  |  | 0.297 |  |  | 0.573 |
| ***Anxiety symptoms*** |  |  |  |  |  |  |  |  |  |
| Anxiety |  |  | 0.791 |  |  | 0.967 |  |  | 0.838 |
| Worrying control |  |  | 0.942 |  |  | 0.967 |  |  | 0.824 |
| Generalised worrying |  |  | 0.942 |  |  | 0.967 |  |  | 0.824 |
| Lack of relaxation |  |  | 0.942 |  |  | 0.967 |  |  | 0.824 |
| Restlessness^a^ |  |  |  |  |  |  |  |  |  |
| Irritability |  |  | 0.069 |  |  | 0.22 |  |  | 0.573 |
| Foreboding |  |  | 0.942 |  |  | 0.942 |  |  | 0.573 |
| ***Depressive symptom score*** | **β (SE)** | **P** | **Q** | **β (SE)** | **P** | **Q** | **β (SE)** | **P** | **Q** |
| Psychological symptoms |  |  | 0.992 |  |  | 0.967 |  |  | 0.776 |
| Somatic symptoms |  |  | 0.027 |  |  | 0.176 |  |  | 0.573 |

*Note*: Estimates and p-values of blank cells are the same as in Supplementary Table 7. ^a^Restlessness symptom was removed from anxiety as duplicate based on the same item from IDS-SR30 used to index Restlessness among psychomotor changes (see Supplementary Table1 for explanation).

##### Supplementary Table 9. Extended NESDA Association Results between IL-6 and Depressive/Anxiety Symptoms

|  |  | **Model 1** |  |  | **Model 2** |  |  | **Model 3** |  |
| --- | --- | --- | --- | --- | --- | --- | --- | --- | --- |
| **Symptom** | **OR (95% CI)** | **P** | **Q** | **OR (95% CI)** | **P** | **Q** | **OR (95% CI)** | **P** | **Q** |
| ***Depressive symptoms*** |  |  |  |  |  |  |  |  |  |
| Anhedonia | 1.39 (1.2-1.62) | <0.001 | <0.001 | 1.3 (1.12-1.52) | <0.001 | 0.009 | 1.27 (1.09-1.49) | 0.002 | 0.026 |
| Depressed mood | 1.21 (1.07-1.37) | 0.002 | 0.007 | 1.15 (1.02-1.3) | 0.022 | 0.08 | 1.15 (1.02-1.3) | 0.024 | 0.106 |
| *Sleeping problems* |  |  |  |  |  |  |  |  |  |
| Falling Asleep | 1.11 (0.99-1.25) | 0.087 | 0.192 | 1.05 (0.94-1.19) | 0.38 | 0.7 | 1.05 (0.93-1.19) | 0.399 | 0.733 |
| During the Night | 1.01 (0.93-1.09) | 0.894 | 0.937 | 1.01 (0.93-1.1) | 0.792 | 0.933 | 1 (0.92-1.09) | 0.945 | 0.982 |
| Waking Up Too Early | 0.97 (0.86-1.09) | 0.59 | 0.721 | 0.96 (0.85-1.08) | 0.468 | 0.7 | 0.97 (0.86-1.09) | 0.608 | 0.786 |
| Sleeping Too Much | 1.35 (1.14-1.59) | <0.001 | 0.002 | 1.26 (1.07-1.49) | 0.006 | 0.032 | 1.25 (1.05-1.48) | 0.011 | 0.06 |
| Fatigue | 1.28 (1.15-1.42) | <0.001 | <0.001 | 1.19 (1.07-1.33) | 0.001 | 0.009 | 1.17 (1.05-1.3) | 0.005 | 0.037 |
| *Appetite changes* |  |  |  |  |  |  |  |  |  |
| Decreased | 1.59 (1.29-1.95) | <0.001 | <0.001 | 1.45 (1.18-1.79) | <0.001 | 0.009 | 1.55 (1.26-1.91) | <0.001 | <0.001 |
| Increased | 1.09 (0.93-1.28) | 0.279 | 0.472 | 1.07 (0.91-1.26) | 0.401 | 0.7 | 0.89 (0.75-1.05) | 0.151 | 0.369 |
| Feelings of inadequacy | 1.08 (0.96-1.21) | 0.228 | 0.418 | 1.04 (0.92-1.17) | 0.513 | 0.706 | 1.04 (0.93-1.18) | 0.48 | 0.733 |
| Cognitive problems | 1.09 (0.95-1.24) | 0.218 | 0.418 | 1.05 (0.92-1.2) | 0.477 | 0.7 | 1.04 (0.91-1.19) | 0.564 | 0.776 |
| *Psychomotor changes* |  |  |  |  |  |  |  |  |  |
| Feeling slowed down | 1.14 (1-1.29) | 0.051 | 0.141 | 1.1 (0.97-1.25) | 0.15 | 0.388 | 1.07 (0.94-1.22) | 0.285 | 0.627 |
| Feeling restless | 1.01 (0.9-1.12) | 0.894 | 0.937 | 0.99 (0.89-1.1) | 0.874 | 0.933 | 1 (0.9-1.12) | 0.945 | 0.982 |
| Suicidal ideation | 0.96 (0.83-1.1) | 0.537 | 0.721 | 0.93 (0.8-1.07) | 0.309 | 0.681 | 0.94 (0.81-1.09) | 0.409 | 0.733 |
| ***Anxiety symptoms*** |  |  |  |  |  |  |  |  |  |
| Anxiety | 1.06 (0.95-1.18) | 0.322 | 0.507 | 1.01 (0.9-1.13) | 0.904 | 0.933 | 1 (0.89-1.12) | 0.982 | 0.982 |
| Worrying control | 1 (0.89-1.13) | 0.999 | 0.999 | 0.97 (0.86-1.09) | 0.605 | 0.783 | 0.96 (0.85-1.08) | 0.5 | 0.733 |
| Generalised worrying | 1.03 (0.92-1.15) | 0.633 | 0.733 | 1 (0.89-1.12) | 0.933 | 0.933 | 1 (0.89-1.12) | 0.976 | 0.982 |
| Lack of relaxation | 1.1 (0.99-1.22) | 0.086 | 0.192 | 1.05 (0.94-1.16) | 0.415 | 0.7 | 1.04 (0.94-1.16) | 0.443 | 0.733 |
| Restlessness^a^ |  |  |  |  |  |  |  |  |  |
| Irritability | 1.05 (0.93-1.18) | 0.452 | 0.663 | 1.01 (0.9-1.14) | 0.873 | 0.933 | 0.99 (0.88-1.12) | 0.875 | 0.982 |
| Foreboding | 0.97 (0.86-1.08) | 0.557 | 0.721 | 0.92 (0.82-1.03) | 0.159 | 0.388 | 0.91 (0.81-1.03) | 0.123 | 0.338 |
| ***Depressive symptom score*** | **β (SE)** | **P** | **Q** | **β (SE)** | **P** | **Q** | **β (SE)** | **P** | **Q** |
| Psychological symptoms | 0.034 (0.012) | 0.006 | 0.018 | 0.024 (0.012) | 0.048 | 0.152 | 0.024 (0.012) | 0.051 | 0.186 |
| Somatic symptoms | 0.048 (0.015) | 0.001 | 0.006 | 0.034 (0.015) | 0.02 | 0.08 | 0.027 (0.015) | 0.067 | 0.209 |

*Note*: ^a^Restlessness symptom was removed from anxiety as duplicate based on the same item from IDS-SR30 used to index Restlessness among psychomotor changes (see Supplementary Table1 for explanation).

#### Results for Mendelian randomization analyses

##### Supplementary Table 10. MR IVW 2-Sample MR Estimates

|  | **CRP (Georgakis)** | | | **CRP (CCGC)** | | | **IL-6 (Georgakis)** | | | **IL-6 (Swerdlow)** | | | **IL-6 (Sarwar)** | | |
| --- | --- | --- | --- | --- | --- | --- | --- | --- | --- | --- | --- | --- | --- | --- | --- |
| **Outcome** | **Estimate (SE)** | **P** | **Q** | **Estimate (SE)** | **P** | **Q** | **Estimate (SE)** | **P** | **Q** | **Estimate (SE)** | **P** | **Q** | **Estimate (SE)** | **P** | **Q** |
| ***Depressive symptoms*** |  |  |  |  |  |  |  |  |  |  |  |  |  |  |  |
| Anhedonia | -0.085 (0.041) | 0.04 | 0.089 | -0.075 (0.081) | 0.356 | 0.473 | 0.046 (0.086) | 0.597 | 0.723 | 0.066 (0.091) | 0.47 | 0.612 | 0.011 (0.037) | 0.771 | 0.867 |
| Depressed mood | -0.074 (0.033) | 0.022 | 0.067 | -0.08 (0.062) | 0.201 | 0.467 | 0.111 (0.081) | 0.172 | 0.295 | -0.061 (0.08) | 0.444 | 0.612 | -0.018 (0.035) | 0.605 | 0.864 |
| Sleeping problems | -0.058 (0.027) | 0.034 | 0.086 | -0.072 (0.04) | 0.07 | 0.314 | 0.191 (0.068) | 0.005 | 0.045 | -0.219 (0.067) | 0.001 | 0.019 | -0.056 (0.03) | 0.057 | 0.651 |
| Fatigue | -0.043 (0.027) | 0.112 | 0.155 | -0.049 (0.039) | 0.208 | 0.467 | 0.245 (0.08) | 0.002 | 0.04 | -0.177 (0.067) | 0.008 | 0.069 | -0.053 (0.029) | 0.072 | 0.651 |
| Appetite changes | -0.044 (0.035) | 0.209 | 0.209 | -0.035 (0.092) | 0.705 | 0.705 | 0.162 (0.113) | 0.151 | 0.295 | -0.118 (0.087) | 0.174 | 0.521 | -0.04 (0.038) | 0.299 | 0.864 |
| Feelings of inadequacy | -0.081 (0.035) | 0.021 | 0.067 | -0.09 (0.1) | 0.367 | 0.473 | 0.045 (0.086) | 0.602 | 0.723 | 0.062 (0.084) | 0.461 | 0.612 | 0.016 (0.037) | 0.667 | 0.864 |
| Cognitive problems | -0.102 (0.035) | 0.004 | 0.023 | -0.126 (0.051) | 0.013 | 0.115 | 0.126 (0.088) | 0.15 | 0.295 | -0.051 (0.086) | 0.552 | 0.621 | -0.018 (0.038) | 0.635 | 0.864 |
| Psychomotor changes | -0.113 (0.058) | 0.054 | 0.107 | -0.161 (0.084) | 0.056 | 0.314 | -0.019 (0.22) | 0.93 | 0.93 | 0.151 (0.143) | 0.291 | 0.582 | 0.053 (0.063) | 0.404 | 0.864 |
| Suicidal ideation | -0.247 (0.083) | 0.003 | 0.023 | -0.245 (0.097) | 0.012 | 0.115 | 0.349 (0.169) | 0.038 | 0.229 | -0.255 (0.166) | 0.123 | 0.521 | -0.065 (0.073) | 0.371 | 0.864 |
| ***Anxiety symptoms*** |  |  |  |  |  |  |  |  |  |  |  |  |  |  |  |
| Anxiety | -0.074 (0.032) | 0.022 | 0.067 | -0.058 (0.062) | 0.351 | 0.473 | 0.101 (0.075) | 0.18 | 0.295 | -0.091 (0.074) | 0.218 | 0.561 | -0.018 (0.033) | 0.575 | 0.864 |
| Worrying control | -0.053 (0.032) | 0.094 | 0.154 | -0.039 (0.046) | 0.396 | 0.475 | 0.109 (0.08) | 0.172 | 0.295 | -0.094 (0.086) | 0.272 | 0.582 | -0.014 (0.035) | 0.682 | 0.864 |
| Generalized worrying | -0.043 (0.029) | 0.142 | 0.171 | -0.047 (0.042) | 0.263 | 0.473 | 0.051 (0.073) | 0.484 | 0.67 | -0.074 (0.077) | 0.335 | 0.603 | -0.006 (0.032) | 0.849 | 0.878 |
| Lack of relaxation | -0.04 (0.03) | 0.18 | 0.192 | -0.023 (0.043) | 0.6 | 0.675 | -0.055 (0.075) | 0.468 | 0.67 | 0.052 (0.086) | 0.548 | 0.621 | 0.022 (0.033) | 0.499 | 0.864 |
| Restlessness | -0.077 (0.043) | 0.071 | 0.128 | -0.042 (0.093) | 0.654 | 0.692 | 0.025 (0.161) | 0.879 | 0.93 | -0.093 (0.13) | 0.476 | 0.612 | -0.02 (0.045) | 0.653 | 0.864 |
| Irritability | -0.049 (0.03) | 0.107 | 0.155 | -0.062 (0.044) | 0.155 | 0.467 | 0.124 (0.076) | 0.103 | 0.295 | -0.107 (0.075) | 0.152 | 0.521 | -0.028 (0.033) | 0.4 | 0.864 |
| Foreboding | -0.056 (0.036) | 0.121 | 0.155 | -0.061 (0.052) | 0.246 | 0.473 | -0.01 (0.091) | 0.912 | 0.93 | 0.001 (0.107) | 0.996 | 0.996 | 0.014 (0.039) | 0.72 | 0.864 |
| ***Depressive symptom score*** |  |  |  |  |  |  |  |  |  |  |  |  |  |  |  |
| Psychological symptoms | -0.071 (0.024) | 0.003 | 0.023 | -0.075 (0.055) | 0.173 | 0.467 | 0.09 (0.055) | 0.099 | 0.295 | -0.004 (0.054) | 0.933 | 0.988 | -0.004 (0.024) | 0.878 | 0.878 |
| Somatic symptoms | -0.045 (0.034) | 0.181 | 0.192 | -0.043 (0.048) | 0.368 | 0.473 | 0.169 (0.106) | 0.111 | 0.295 | -0.141 (0.083) | 0.088 | 0.521 | -0.036 (0.034) | 0.289 | 0.864 |

##### Supplementary Table 11. MR IVW 1-Sample MR Estimates

|  | **CRP (Georgakis)** | | | **CRP (CCGC)** | | | **IL-6 (Georgakis)** | | | **IL-6 (Swerdlow)** | | | **IL-6 (Sarwar)** | | |
| --- | --- | --- | --- | --- | --- | --- | --- | --- | --- | --- | --- | --- | --- | --- | --- |
| **Outcome** | **Estimate (SE)** | **P** | **Q** | **Estimate (SE)** | **P** | **Q** | **Estimate (SE)** | **P** | **Q** | **Estimate (SE)** | **P** | **Q** | **Estimate (SE)** | **P** | **Q** |
| ***Depressive symptoms*** |  |  |  |  |  |  |  |  |  |  |  |  |  |  |  |
| Anhedonia | -0.112 (0.044) | 0.011 | 0.053 | -0.062 (0.056) | 0.271 | 0.383 | 0.005 (0.082) | 0.948 | 0.973 | -0.051 (0.095) | 0.588 | 0.705 | -0.033 (0.113) | 0.771 | 0.867 |
| Depressed mood | -0.078 (0.036) | 0.032 | 0.093 | -0.059 (0.046) | 0.203 | 0.365 | 0.092 (0.078) | 0.234 | 0.421 | 0.067 (0.08) | 0.398 | 0.597 | 0.055 (0.107) | 0.605 | 0.864 |
| Sleeping problems | -0.068 (0.03) | 0.026 | 0.093 | -0.072 (0.033) | 0.027 | 0.216 | 0.169 (0.067) | 0.012 | 0.11 | 0.217 (0.066) | 0.001 | 0.019 | 0.17 (0.089) | 0.057 | 0.651 |
| Fatigue | -0.044 (0.03) | 0.145 | 0.217 | -0.031 (0.033) | 0.337 | 0.433 | 0.235 (0.076) | 0.002 | 0.034 | 0.179 (0.066) | 0.007 | 0.062 | 0.159 (0.089) | 0.072 | 0.651 |
| Appetite changes | -0.034 (0.039) | 0.389 | 0.389 | -0.049 (0.066) | 0.458 | 0.485 | 0.151 (0.108) | 0.164 | 0.363 | 0.124 (0.086) | 0.15 | 0.45 | 0.12 (0.116) | 0.299 | 0.864 |
| Feelings of inadequacy | -0.097 (0.039) | 0.012 | 0.053 | -0.058 (0.074) | 0.43 | 0.484 | 0.031 (0.082) | 0.703 | 0.904 | -0.061 (0.084) | 0.463 | 0.641 | -0.048 (0.112) | 0.667 | 0.864 |
| Cognitive problems | -0.119 (0.039) | 0.002 | 0.042 | -0.111 (0.042) | 0.008 | 0.15 | 0.095 (0.084) | 0.257 | 0.421 | 0.055 (0.086) | 0.524 | 0.674 | 0.055 (0.115) | 0.635 | 0.864 |
| Psychomotor changes | -0.116 (0.065) | 0.076 | 0.152 | -0.115 (0.07) | 0.103 | 0.356 | -0.022 (0.21) | 0.917 | 0.973 | -0.137 (0.142) | 0.334 | 0.597 | -0.159 (0.19) | 0.404 | 0.864 |
| Suicidal ideation | -0.209 (0.101) | 0.038 | 0.093 | -0.15 (0.112) | 0.178 | 0.356 | 0.324 (0.161) | 0.044 | 0.262 | 0.255 (0.165) | 0.122 | 0.45 | 0.198 (0.221) | 0.371 | 0.864 |
| ***Anxiety symptoms*** |  |  |  |  |  |  |  |  |  |  |  |  |  |  |  |
| Anxiety | -0.075 (0.037) | 0.041 | 0.093 | -0.066 (0.045) | 0.141 | 0.356 | 0.096 (0.072) | 0.182 | 0.363 | 0.096 (0.074) | 0.193 | 0.497 | 0.055 (0.099) | 0.575 | 0.864 |
| Worrying control | -0.05 (0.036) | 0.158 | 0.219 | -0.052 (0.038) | 0.171 | 0.356 | 0.117 (0.076) | 0.125 | 0.363 | 0.099 (0.081) | 0.222 | 0.5 | 0.043 (0.104) | 0.682 | 0.864 |
| Generalized worrying | -0.043 (0.033) | 0.184 | 0.23 | -0.052 (0.035) | 0.143 | 0.356 | 0.031 (0.07) | 0.651 | 0.901 | 0.077 (0.074) | 0.296 | 0.593 | 0.018 (0.096) | 0.849 | 0.878 |
| Lack of relaxation | -0.058 (0.034) | 0.084 | 0.152 | -0.039 (0.036) | 0.276 | 0.383 | -0.039 (0.072) | 0.591 | 0.886 | -0.041 (0.088) | 0.644 | 0.724 | -0.067 (0.099) | 0.499 | 0.864 |
| Restlessness | -0.054 (0.05) | 0.279 | 0.314 | -0.024 (0.066) | 0.711 | 0.711 | 0.005 (0.153) | 0.973 | 0.973 | 0.105 (0.124) | 0.396 | 0.597 | 0.061 (0.135) | 0.653 | 0.864 |
| Irritability | -0.044 (0.034) | 0.192 | 0.23 | -0.077 (0.037) | 0.036 | 0.216 | 0.135 (0.072) | 0.062 | 0.279 | 0.114 (0.074) | 0.126 | 0.45 | 0.084 (0.1) | 0.4 | 0.864 |
| Foreboding | -0.061 (0.041) | 0.135 | 0.217 | -0.062 (0.044) | 0.156 | 0.356 | 0.004 (0.087) | 0.96 | 0.973 | 0.01 (0.106) | 0.927 | 0.927 | -0.043 (0.119) | 0.72 | 0.864 |
| ***Depressive symptom score*** |  |  |  |  |  |  |  |  |  |  |  |  |  |  |  |
| Psychological symptoms | -0.074 (0.027) | 0.006 | 0.053 | -0.051 (0.043) | 0.239 | 0.383 | 0.072 (0.052) | 0.166 | 0.363 | 0.009 (0.053) | 0.863 | 0.914 | 0.011 (0.071) | 0.878 | 0.878 |
| Somatic symptoms | -0.038 (0.038) | 0.318 | 0.336 | -0.03 (0.038) | 0.43 | 0.484 | 0.157 (0.102) | 0.125 | 0.363 | 0.147 (0.077) | 0.056 | 0.334 | 0.109 (0.103) | 0.289 | 0.864 |

##### Supplementary Table 12. MR IVW 2-Sample MR Heterogeneity Estimates

|  |  | **CRP (Georgakis)** | |  | **CRP (CCGC)** | |  | **IL-6 (Georgakis)** | |  | **IL-6 (Swerdlow)** | |
| --- | --- | --- | --- | --- | --- | --- | --- | --- | --- | --- | --- | --- |
| **Outcome** |  | **Q** | **P** |  | **Q** | **P** |  | **Q** | **P** |  | **Q** | **P** |
| ***Depressive symptoms*** |  |  |  |  |  |  |  |  |  |  |  |  |
| Anhedonia |  | 32.76 | 0.085 |  | 5.33 | 0.07 |  | 3.93 | 0.686 |  | 2.28 | 0.319 |
| Depressed mood |  | 21.11 | 0.574 |  | 3.53 | 0.171 |  | 1.98 | 0.922 |  | 1 | 0.605 |
| Sleeping problems |  | 13.93 | 0.929 |  | 2.07 | 0.356 |  | 5.37 | 0.498 |  | 1.32 | 0.518 |
| Fatigue |  | 14.33 | 0.917 |  | 1.24 | 0.537 |  | 8.4 | 0.21 |  | 0.54 | 0.762 |
| Appetite changes |  | 22.16 | 0.51 |  | 6.51 | 0.038 |  | 9.77 | 0.135 |  | 0.53 | 0.769 |
| Feelings of inadequacy |  | 24.39 | 0.382 |  | 8.14 | 0.017 |  | 1.69 | 0.946 |  | 0.01 | 0.995 |
| Cognitive problems |  | 16.66 | 0.825 |  | 0.58 | 0.749 |  | 4.84 | 0.565 |  | 0.24 | 0.885 |
| Psychomotor changes |  | 17.41 | 0.788 |  | 0.13 | 0.938 |  | 13.77 | 0.032 |  | 1.27 | 0.53 |
| Suicidal ideation |  | 35.37 | 0.048 |  | 2.02 | 0.363 |  | 2.61 | 0.856 |  | 0.02 | 0.99 |
| ***Anxiety symptoms*** |  |  |  |  |  |  |  |  |  |  |  |  |
| Anxiety |  | 26.36 | 0.284 |  | 4.08 | 0.13 |  | 0.91 | 0.989 |  | 1.06 | 0.589 |
| Worrying control |  | 9.69 | 0.993 |  | 1.29 | 0.524 |  | 2.89 | 0.823 |  | 2.38 | 0.304 |
| Generalized worrying |  | 13.75 | 0.934 |  | 0.36 | 0.833 |  | 4.43 | 0.618 |  | 2.27 | 0.321 |
| Lack of relaxation |  | 11.52 | 0.977 |  | 1.92 | 0.383 |  | 3.25 | 0.777 |  | 2.71 | 0.258 |
| Restlessness |  | 24.75 | 0.363 |  | 4.92 | 0.085 |  | 14.52 | 0.024 |  | 3.29 | 0.193 |
| Irritability |  | 12.37 | 0.964 |  | 1.32 | 0.516 |  | 3.18 | 0.786 |  | 1.9 | 0.386 |
| Foreboding |  | 16.45 | 0.836 |  | 1.25 | 0.535 |  | 3.45 | 0.75 |  | 2.85 | 0.241 |
| **Depressive symptom score** |  |  |  |  |  |  |  |  |  |  |  |  |
| Psychological symptoms |  | 26.96 | 0.258 |  | 6.08 | 0.048 |  | 3.52 | 0.742 |  | 0.8 | 0.669 |
| Somatic symptoms |  | 26.05 | 0.299 |  | 2.2 | 0.334 |  | 10.93 | 0.09 |  | 2.28 | 0.32 |

##### Supplementary Table 13. MR IVW 1-Sample MR Heterogeneity Estimates

|  |  | **CRP (Georgakis)** | |  | **CRP (CCGC)** | |  | **IL-6 (Georgakis)** | |  | **IL-6 (Swerdlow)** | |
| --- | --- | --- | --- | --- | --- | --- | --- | --- | --- | --- | --- | --- |
| **Outcome** |  | **Q** | **P** |  | **Q** | **P** |  | **Q** | **P** |  | **Q** | **P** |
| ***Depressive symptoms*** |  |  |  |  |  |  |  |  |  |  |  |  |
| Anhedonia |  | 30.34 | 0.14 |  | 5.54 | 0.136 |  | 4.2 | 0.649 |  | 2.51 | 0.285 |
| Depressed mood |  | 21.73 | 0.536 |  | 4.17 | 0.243 |  | 2.43 | 0.876 |  | 0.88 | 0.645 |
| Sleeping problems |  | 13.5 | 0.94 |  | 2.22 | 0.527 |  | 6.47 | 0.373 |  | 1.33 | 0.514 |
| Fatigue |  | 14.73 | 0.904 |  | 1.99 | 0.575 |  | 8.24 | 0.221 |  | 0.34 | 0.843 |
| Appetite changes |  | 23 | 0.461 |  | 7.16 | 0.067 |  | 9.92 | 0.128 |  | 0.31 | 0.858 |
| Feelings of inadequacy |  | 23.51 | 0.431 |  | 9.57 | 0.023 |  | 1.82 | 0.936 |  | 0.02 | 0.992 |
| Cognitive problems |  | 15.77 | 0.865 |  | 0.75 | 0.862 |  | 5.62 | 0.467 |  | 0.19 | 0.909 |
| Psychomotor changes |  | 17.99 | 0.758 |  | 1.13 | 0.771 |  | 13.77 | 0.032 |  | 1.45 | 0.484 |
| Suicidal ideation |  | 41.09 | 0.012 |  | 5.66 | 0.129 |  | 2.84 | 0.828 |  | 0 | 0.998 |
| ***Anxiety symptoms*** |  |  |  |  |  |  |  |  |  |  |  |  |
| Anxiety |  | 27.43 | 0.238 |  | 4.56 | 0.207 |  | 0.92 | 0.988 |  | 0.88 | 0.643 |
| Worrying control |  | 10.5 | 0.988 |  | 1.95 | 0.583 |  | 2.4 | 0.879 |  | 2.19 | 0.335 |
| Generalized worrying |  | 14.14 | 0.923 |  | 0.53 | 0.913 |  | 4.72 | 0.58 |  | 2.15 | 0.341 |
| Lack of relaxation |  | 10.34 | 0.989 |  | 2.85 | 0.416 |  | 3.48 | 0.746 |  | 2.88 | 0.236 |
| Restlessness |  | 26.88 | 0.261 |  | 5.23 | 0.156 |  | 14.57 | 0.024 |  | 3.03 | 0.219 |
| Irritability |  | 13.26 | 0.946 |  | 2 | 0.573 |  | 2.35 | 0.884 |  | 1.6 | 0.448 |
| Foreboding |  | 16.62 | 0.828 |  | 1.34 | 0.721 |  | 3.46 | 0.749 |  | 2.84 | 0.242 |
| **Depressive symptom score** |  |  |  |  |  |  |  |  |  |  |  |  |
| Psychological symptoms |  | 28.33 | 0.204 |  | 8.07 | 0.045 |  | 4.32 | 0.633 |  | 0.78 | 0.677 |
| Somatic symptoms |  | 26.9 | 0.26 |  | 2.46 | 0.482 |  | 11.17 | 0.083 |  | 1.93 | 0.381 |

### eDISCUSSION

##### Supplementary Table 14. Overview on result consistency of associations between inflammation and symptoms

|  |  |  | **MR (UKB)** | |
| --- | --- | --- | --- | --- |
| **Symptom** | **CRP (pooled)** | **IL-6 (NESDA)** | **CRP** | **IL-6** |
| ***Depressive symptoms*** |  |  |  |  |
| Fatigue | ↑ | ↑ |  | ↑ |
| Sleeping problems | ↑ |  |  | ↑ |
| Insomnia (NESDA) |  |  | NA | NA |
| Hypersomnia (NESDA) | ↑ | ↑ | NA | NA |
| Appetite changes | ↑ |  |  |  |
| increase (NESDA) | ↑ |  | NA | NA |
| decrease (NESDA) |  | ↑ | NA | NA |
| Depressed mood | ↑ | ↑ |  |  |
| Anhedonia |  | ↑ |  |  |
| Suicidality |  |  | ↓ |  |
| Psychomotor changes |  |  |  |  |
| Agitation (NESDA) |  |  | NA | NA |
| Retardation (NESDA) |  |  | NA | NA |
| Cognitive problems |  |  | ↓ |  |
| Feelings of inadequacy |  |  |  |  |
| ***Anxiety symptoms*** |  |  |  |  |
| Irritability | ↑ |  |  |  |
| Worrying control | ↑ |  |  |  |
| Generalised worrying |  |  |  |  |
| Lack of relaxation |  |  |  |  |
| Anxiety |  |  |  |  |
| Restlessness |  |  |  |  |
| Foreboding |  |  |  |  |
| ***Depressive symptom scores*** |  |  |  |  |
| Psychological |  |  | ↓ |  |
| Somatic |  |  |  |  |

*Note*: ↑=FDR-corrected increase in phenotype; ↓=FDR-corrected decrease in phenotype; *=granular symptom results are not based on pooled estimates, but only on NESDA based on availability; NA=not applicable. Phenotypic results are based on significance of model 2. MR results are based on Georgakis *et al.*[17] instruments for CRP and IL-6 and all significant results are shown if at least one of the 1- and 2-sample MR estimates was FDR-corrected significant.

##### Supplementary Figure 3. Hypothesised model outlining the complex relationship between inflammation, BMI and depression

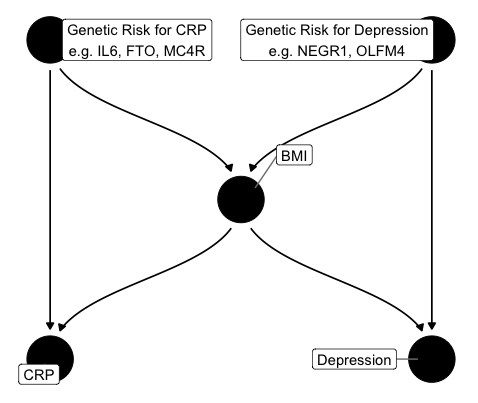
